# Cerebral hemodynamics in early adolescents with congenital heart disease after infant open-heart surgery

**DOI:** 10.1101/2025.10.27.25338861

**Authors:** François Mojon, Beatrice Latal, Raimund Kottke, Oliver Kretschmar, Ruth Tuura O’Gorman, Melanie Ehrler

**Affiliations:** Child Development Center, University Children’s Hospital Zurich, Zurich, Switzerland; Children’s Research Center, University Children’s Hospital Zurich, Zurich, Switzerland; URPP Adaptive Brain Circuits in Development and Learning, University of Zurich; Department of Diagnostic Imaging, University Children’s Hospital Zurich, Zurich, Switzerland; Department Pediatric Cardiology, Pediatric Heart Center, University Children’s Hospital Zurich, Zurich, Switzerland; Center for MR Research, University Children’s Hospital Zurich, Zurich, Switzerland; Department of Forensic and Neurodevelopmental Sciences, Institute of Psychiatry, Psychology, and Neuroscience, King’s College London, UK

**Author notes:** shared last authorship (Ruth Tuura O’Gorman, Melanie Ehrler). **Corresponding Author Info:** François Mojon, Child Development Center, University Children’s Hospital Zurich, Lenggstrasse 30, Zurich 8008, Switzerland.

**Keywords:** congenital heart disease, cerebral blood flow, cerebral hemodynamics, phase-contrast magnetic resonance imaging, executive functions, adolescents

## Abstract

Fetuses and infants with congenital heart disease (CHD) show altered cerebral hemodynamics. However, little is known about whether those alterations persist throughout childhood. We assessed cerebral hemodynamics in 34 adolescents with CHD aged 10–15 years (35% females), who had infant open-heart surgery, and 49 healthy controls (51% females), and investigated the relationship between cerebral hemodynamics and executive functions. Using phase-contrast magnetic resonance imaging (pcMRI) we measured the average and peak blood velocity and average blood flow in the bilateral internal carotid arteries (ICA) and the average blood velocity and flow in the internal jugular veins (IJV). ICA blood flow was used to calculate the indexed cerebral blood flow (iCBF) adjusted for total brain volume. Vertebral arteries (VA) were analyzed post-hoc using a vessel angle correction. Cerebral hemodynamic parameters were compared between patients and healthy controls and correlated with an executive function summary score. Differences in anterior hemodynamic parameters between patients with CHD vs. healthy controls did not survive FDR correction (all p>0.1, β:0.004–0.297), but lower blood velocity and flow in the left VA in patients with CHD remained significant after FDR-correction (both corrected p=0.044, β=0.248–0.314). Hemodynamic parameters were not significantly associated with the executive function summary score (all p>0.8, β:0.023–0.119). Overall, patients with CHD who underwent infant open-heart surgery have widely preserved anterior cerebral hemodynamics during adolescence, but demonstrate alterations in the posterior circulation. Further, hemodynamic parameters show no association with executive functions and thus, may not be a risk factor for executive function impairments.

**Highlights:**

- Adolescents with CHD have widely preserved anterior cerebral hemodynamics
- Posterior cerebral perfusion might be reduced compared to the healthy population
- No association between cerebral hemodynamics and executive function performance

## 1. Introduction

Affecting approximately 8 in 1000 live births, congenital heart disease (CHD) is the most common congenital defect (Bernier et al., 2010). Thanks to improvements in diagnostic and therapeutic procedures, the population of patients with CHD has been constantly growing and the number of adults with CHD has surpassed that of children with CHD (Marelli et al., 2014). Consequently, the focus of research and patient care has been increasingly shifted towards long-term outcome and comorbidities of patients with CHD (Ruperti-Repilado et al., 2021). Impaired neurodevelopmental outcomes constitute the most frequent non-cardiac comorbidities in patients with CHD (Cassidy et al., 2018; Feldmann et al., 2021). Among these, deficits in higher-order cognitive functions, such as in executive functions, are found in patients with CHD, and have been shown to contribute substantially to long-term developmental burden (Spillmann et al., 2023). Executive functions encompass advanced neurocognitive abilities, such as working memory, inhibition, cognitive flexibility, planning, and fluency (Diamond, 2013). The impact of CHD on the brain is multifactorial (Howell et al., 2019; Wernovsky, 2006). Although poorly investigated, altered cerebral hemodynamics and subsequent restricted blood delivery to the brain might contribute to the persisting neurologic sequelae in patients with CHD (Cheng et al., 2014; De Silvestro et al., 2023; Wernovsky, 2006). Previous studies suggest that patients with CHD show alterations of cerebral hemodynamics preoperatively. In fetuses with CHD, a mechanism called brain sparing is thought to enhance cerebral blood flow, via a reduced cerebroplacental resistance ratio, in comparison to that in healthy controls (Donofrio et al., 2003). According to a recent review by De Silvestro et al. (2023), alterations of cerebral hemodynamics, assessed by various neuroimaging techniques, persist after birth in infants (i.e. first year of life) with severe CHD (De Silvestro et al., 2023). Neuroimaging techniques in the review included doppler ultrasound, phase-contrast magnetic resonance imaging (pcMRI) and arterial spin labeling (ASL). However, heterogeneous results were obtained when comparing cerebral hemodynamic parameters between patients with CHD and healthy controls or literature reference values for healthy infants. While some studies reported altered values in infants with CHD, such as lower blood flow or altered blood velocities (e.g., lower average velocity or higher pulsatility index), others did not observe significant differences between patients with CHD and healthy infants. In general, alterations of cerebral hemodynamics, if observed, were more frequent in the most severe cases of CHD, such as single ventricle and cyanotic heart defects, possibly due to the systemic-to-pulmonary shunt steal phenomenon.

However, little is known about whether cerebral hemodynamic alterations persist beyond the presurgical period and whether these alterations may underly long-term neurodevelopmental sequelae. To date, only two studies have investigated if alterations of cerebral hemodynamics remain throughout childhood. Schmithorst et al. (2022) studied regional cerebral perfusion using ASL in a cohort of patients with mixed CHD, aged 6–21 years, and its relationship with neurodevelopmental outcome. Reduced regional cerebral perfusion in frontal, parietal and subcortical regions, coupled to a steeper age-related decline of regional cerebral perfusion in late adolescence, was found in patients with CHD compared to healthy controls. The authors further provide first evidence that reduced regional cerebral perfusion is associated with lower domain-specific cognitive performance in children and adolescents with CHD (Schmithorst et al., 2022). Easson et al. (2023) assessed global gray matter and regional gray matter perfusion using ASL in a cohort of patients with complex CHD, aged 16–24 years. They observed reduced global and regional gray matter perfusion in female patients with CHD compared to female controls, but no differences were found in males (Easson et al., 2023).

In this study, using phase contrast MRI (pcMRI), we assessed arterial and venous cerebral hemodynamics in a cohort of adolescents with CHD who underwent cardiopulmonary bypass surgery within the first year of life. PcMRI measures velocities, such as arterial or venous blood velocity, from the phase change induced by motion along a bipolar gradient (Wymer et al., 2020) and has been shown to quantify cerebral vascular flow reliably (Sakhare et al., 2019). Our primary aim was to compare cerebral hemodynamics between adolescents with CHD and healthy controls and to investigate the correlation between cerebral hemodynamic parameters and executive function performance. As a secondary aim, we compared cerebral hemodynamics between patients with cyanotic (cCHD), acyanotic (aCHD) CHD and healthy controls, since alterations of cerebral hemodynamics have been described to be most pronounced in infants with the most severe cases of CHD, such as cCHD (De Silvestro et al., 2023). We hypothesized that lower cerebral blood velocities and blood flow would be present in patients compared to controls. We further hypothesized that lower cerebral blood flow and velocity would be associated with poorer executive function performance, since previous studies have shown reduced executive functions in patients with CHD and we considered reduced cerebral blood flow a potential contributing risk factor.

## 2. Methods

### 2.1 Study Sample

The data for this study were collected as part of the Teenheart Study (Ehrler et al., 2019), a prospective cohort study carried out at the University Children’s Hospital of Zurich between April 2019 and September 2021, assessing neurocognitive outcomes and cerebral MRI of adolescents with CHD. The Teenheart Study includes patients with complex CHD who underwent cardiopulmonary bypass surgery at the University Children’s Hospital Zurich between July 2004 and July 2012 and before the age of 1 year. Patients were aged between 10 and 15 years at the time of the assessment and lived in Switzerland. Patients with a known genetic comorbidity or dysmorphic syndrome were excluded. Of 178 eligible patients, 100 (56%) participated in the Teenheart Study. Of the 100 participating patients, 60 (60%) underwent a cerebral MRI and in 36 (36%) patients a pcMRI sequence was obtained.

A total of 104 healthy adolescents aged 10–15 years were recruited as a control group. Controls were term-born (>36 weeks of gestation), and had no known neurological, congenital, or developmental disorder. Of the 104 controls, 96 (92%) underwent a cerebral MRI and in 55 (53%) controls a pcMRI sequence was obtained.

The detailed recruitment procedure is shown in the supplementary Figure 1. Written informed consent was obtained from all the participants older than 14 years of age and from the participant’s legal guardian. The ethics committee of the Canton of Zurich, Switzerland approved the study (KEK 2019–00035).

### 2.2 Phase-contrast magnetic resonance imaging (pcMRI)

MRI of the brain was performed on a 3T GE MR750 MRI scanner (GE Healthcare, Milwaukee, WI). Safety screening questionnaires were previously filled out by the parents. Study participants were awake, were monitored by pulse oximetry and hearing protection was provided during the whole scanning procedure. Axial phase-contrast images were acquired using a single-slice sequence near the base of the cerebellum, perpendicular to the internal carotid arteries and internal jugular veins, as visualized on a sagittal and coronal reformat of a three-dimensional T1-weighted scan (Supplementary Figure 2). Cardiac gating was obtained using a peripheral pulse transducer. The following scanning parameters were applied: field of view (FOV)=180mm, repetition time/echo time (TR/TE)=7.50/3.95ms, slice thickness=4mm, maximal encoding velocity (V_ENC_)=120cm/s, resolution=0.7×0.7mm^2^, flip angle=20°. Furthermore, T1-weighted scans were performed on all study participants (inversion time=600ms, repetition time/echo time [TR/TE]=11.39/5.18ms, resolution=1×1×1mm^3^, flip angle=8°) and reviewed by a neuroradiologist. After pcMRI-acquisition, we excluded 8 participants because of incidental MRI findings (0 patients, 1 control [big cystic lesion]), excessive motion artefacts (2 patients, 3 controls) or disrupted vessel signal due to braces (0 patients, 2 controls), resulting in a total dataset of 34 patients with CHD and 49 healthy controls.

### 2.3 Assessment of cerebral hemodynamics

Measurements of cerebral hemodynamics were conducted by a single blinded rater using a commercial postprocessing tool (Medis QFlow software version 8.1, Medis Medical Imaging Systems). Regions of interest were manually drawn bilaterally around the internal carotid arteries (ICA) and internal jugular veins (IJV), measuring average blood velocity (ABV; cm/s), peak blood velocity (PBV; cm/s; assessed for ICA only) and blood flow (BF; ml/min) (Figure 1). Together, the ICA and the IJV constitute a major part of the anterior cerebral circulation, whereas the vertebral arteries (VA) build the major posterior cerebral arterial blood supply (Charlick and Das, 2024). Vessels that were not perpendicular to the scan were excluded from the measurements (1 unilateral ICA [1 patient, 0 controls], 7 unilateral IJVs [4 patients, 3 controls], 2 bilateral IJVs [0 patients, 2 controls]). In one patient, 1 ICA was further unilaterally excluded due to a hemodynamically relevant anatomical variation (persistent primitive trigeminal artery). The sum of the bilateral ICA BF (2 missing, see above) was further divided by the total brain volume to obtain the indexed cerebral blood flow (iCBF; ml/min/100g). Total brain volume was derived from the T1-weighted images using the FreeSurfer image analysis suite version 7.1 (Fischl, 2012). Table 1 gives an overview of the eleven different cerebral hemodynamic parameters primary assessed in this study. The above-described measurements were further conducted on 10 pcMRI-scans (5 patients, 5 healthy controls) by a second blinded rater to check for intraclass correlation (ICC).

**Figure 1.**
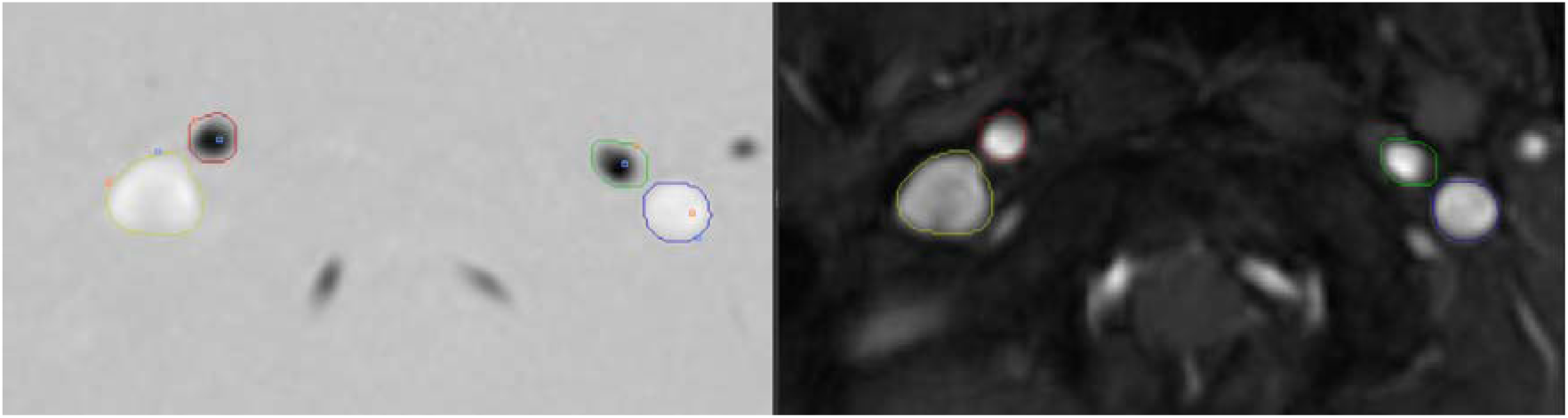
pcMRI and T1-weighted MRI with regions of interest. Left: pcMRI-sequence; right: T1-weighted MRI-sequence. Regions of interest were manually drawn around the right ICA (red), left ICA (green), right IJV (yellow), and left IJV (blue).

**Table 1.** Cerebral hemodynamic parameters primary assessed in this study.

| Parameter | Unit |
| --- | --- |
| Right and left ICA (internal carotid artery) |  |
| ABV (average blood velocity) | cm/s |
| PBV (peak blood velocity) | cm/s |
| BF (blood flow) | ml/min |
| Bilateral ICA (internal carotid arteries) |  |
| iCBF (indexed cerebral blood flow) | ml/min/100g |
| Right and left IJV (internal jugular vein) |  |
| ABV (average blood velocity) | cm/s |
| BF (blood flow) | ml/min |

Due to the suboptimal (non-perpendicular) slice positioning the hemodynamics of the VA were not included in the main analysis, but the velocities (ABV and PBV) and flow values (BF) were included in a post-hoc exploratory analysis after estimating the vessel angle from the ratio of the lengths of the short and long axes of the vessel as it intersects the imaging plane, such as described for doppler ultrasonography in Goffi et al. (2025).

### 2.4 Executive functions performance

An extensive neuropsychological test battery was conducted using standardized tests for working memory, inhibition, cognitive flexibility, fluency, and problem solving (supplementary Table 1). An age-adjusted summary score for executive function performance was computed as described in von Werdt et al. (2024).

### 2.5 Additional patient information

Medical data was prospectively collected from patient’s charts. The sum of the highest maternal and paternal education on a 6-point scale each was used as a proxy for socio-economic status (SES; 1=no high-school degree, 2=high-school degree, 3=apprenticeship, 4=higher diploma for craftsmen or craftswomen, 5=advanced diploma of higher education, 6=university degree). To increase statistical power, missing SES values (3 patients, 5 controls) were substituted by the group-specific mean value.

### 2.6 Statistics

Data was visually checked for normality. Sample characteristics were described using amount and percentage for nominal data, mean and SD (standard deviation) for normally distributed continuous data, or median and range for non-normally distributed continuous or ordinal data. Sample characteristics were compared between patient and control groups as well as between all study participants and participants included in this analysis with two-tailed t-tests for continuous data, Mann-Whitney U tests for ordinal data, and chi-square tests for nominal data.

To investigate group differences in cerebral hemodynamics, separate linear regression models were estimated with cerebral hemodynamic parameters as dependent variables and group (CHD vs. controls) or subgroup (aCHD vs. cCHD vs. controls) as independent variable. Age and sex were included as covariates. Post-hoc, we estimated linear regression models for vertebral hemodynamic parameters comparing CHD vs. controls. We further investigated an interaction post-hoc between group and sex, as this has been previously reported by Easson et al. (2023).

To investigate associations with executive functions, separate linear regression models were estimated with the executive function summary score as dependent variable and cerebral hemodynamic parameters as independent variables. For this analysis, only cerebral hemodynamic parameters were considered which differed significantly between groups or subgroups before FDR-correction. Significant parameters of the post-hoc vertebral analysis were not included. Age, sex, SES and group were included as covariates.

FDR (False Discovery Rate)-correction was applied to adjust for multiple comparisons (Benjamini and Hochberg, 1995) and *p*-values<0.05 were considered significant. All statistical analyses were performed using the R statistical package, version 4.2.3 (R Core Team, 2023).

## 3. Results

### 3.1 Sample characteristics & structural image evaluation

This study includes 34 patients with CHD and 49 healthy controls between 10 and 15 years of age. Table 2 shows the participant demographics and Table 3 the medical characteristics of the CHD group. Patients and controls did not significantly differ in sex, but patients were older and had a lower SES, executive function summary score, IQ and brain volume relative to controls. Out of 34 patients with CHD, 65% had cyanotic and 15% univentricular CHD.

**Table 2.** Sample demographics of patient and control cohort.

| **Variable** | **Patients with CHD**<br>**(n=34)** | **Healthy controls**<br>**(n=49)** | **P-value** | **Effect**<br>**size** |
| --- | --- | --- | --- | --- |
| **Age in years, mean (SD)** | 13.54 (1.06) | 12.92 (1.45) | 0.027<sup>a</sup> | 0.48<sup>d</sup> |
| **Female sex, N (%)** | 12 (35.3) | 25 (51.0) | 0.233<sup>c</sup> | 0.13<sup>e</sup> |
| **SES, median (range)** | 8 (4–12) | 10 (5–12) | 0.002<sup>b</sup> | 0.75<sup>d</sup> |
| **Executive functions**<br>**summary score, mean (SD)** | -1.05 (1.29) | -0.08 (1.06) | <0.001<sup>a</sup> | 0.83<sup>d</sup> |
| **IQ\*, mean (SD)** | 100.74 (12.45) | 110.26 (9.86) | <0.001<sup>a</sup> | 0.86<sup>d</sup> |
| **Brain volume, cm<sup>3</sup>, mean (SD)** | 1121.27 (143.42) | 1207.97 (100.72) | 0.004<sup>a</sup> | 0.72<sup>d</sup> |

Incomplete data for IQ (missing n=2) and SES = Socioeconomic status (missing n=8). <sup>a</sup> two-tailed t-test, <sup>b</sup> Mann-Whitney U tests, <sup>c</sup> chi-square test
<sup>d</sup> Cohen's d, <sup>e</sup> Phi; \*IQ was assessed using the WISC-4 short form, corrected according to Ehrler et al. (2020).
CHD=congenital heart disease.

**Table 3.** Medical characteristics of patient cohort.

| **Variable** | **Patients with CHD (n=34)** |
| --- | --- |
| **Cardiac diagnosis, N (%)** |  |
| Acyanotic CHD | 12 (35) |
| Ventricular septal defect | 4 (12) |
| Atrioventricular septal defect | 3 (9) |
| Aortic coarctation | 2 (6) |
| Atrial septal defect | 1 (3) |
| Pulmonary stenosis | 1 (3) |
| Anomalous left coronary artery from the pulmonary artery | 1 (3) |
| Cyanotic CHD | 22 (65) |
| Transposition of the great arteries | 11 (32) |
| Hypoplastic left heart syndrome | 3 (9) |
| Tetralogy of Fallot | 2 (6) |
| Total anomalous pulmonary venous connection | 2 (6) |
| Truncus arteriosus communis | 2 (6) |
| Tricuspid atresia | 1 (3) |
| Double inlet left ventricle | 1 (3) |
| **Univentricular heart defect, N (%)** | 5 (15) |
| **Gestational age, wk, mean (SD)** | 39.47 (1.76) |
| **Birth weight, g, mean (SD)** | 3317.06 (516.93) |
| **Head circumference at birth, cm, mean (SD)** | 34.62 (1.72) |
| **Prenatal diagnosis, N (%)** | 6 (18) |
| **CHD complexity\*, N (%)** |  |
| Simple | 5 (15) |
| Moderate | 11 (32) |
| Severe | 18 (53) |
| **RACHS score, median (range)** | 3 (2–6) |
| **Age at surgery\*\*, mo, mean (SD)** | 2.15 (2.83) |
| **Mean preoperative saturation\*\*, %, mean (SD)** | 88.55 (9.10) |
| **Lowest temperature\*\*, °C, mean (SD)** | 28.97 (3.49) |
| **ECC time\*\*, min, mean (SD)** | 163.44 (58.28) |
| **Postoperative time in ICU\*\*, d, mean (SD)** | 8.76 (6.81) |
| **Postoperative time of hospitalisation\*\*, d, mean (SD)** | 36.03 (36.47) |
| **ECMO, N (%)** | 2 (6) |
| **Total number of CPB surgeries, median (range)** | 1 (1–3) |
| **Structural brain abnormality, N (%)** | 3 (9) |
| Focal abnormalities | 3 (9) |
| Global anomalies | 0 (0) |
| **Echocardiography\*\*\*** |  |
| Atrioventricular valve insufficiency, N (%) | 1 (3) |

| Decreased cardiac function, N (%) | 1 (3) |
| --- | --- |
| Incomplete data for head circumference (missing n=3), mean preoperative saturation (n=1) and echocardiographic cardiac function (missing n=1). *According to Warnes et al. (2001). **Correspond to the first CPB surgery. ***Information was taken from the last echocardiographic examination before participating in this study.<br>CHD=congenital heart disease; RACHS=risk adjustment for congenital heart surgery; ECC: extracorporeal circulation; ECMO=extracorporeal membrane oxygenation; CPB=cardiopulmonary bypass. |  |

9% of the patients showed structural brain abnormalities in the T_1_-weighted image (2 small focal infarctions without a clinical correlate, 1 focal atrophy). No brain abnormalities were detected in controls with conventional T1-weighted MRI.

Patients included in the analysis of this study did not differ in age (p=0.193, Cohen’s d=0.17), sex (p=0.953, Phi=0.00), or CHD complexity (according to Warnes et al. [2001]; p=0.126, Phi=0.20) compared to study participants who were not included in this analysis due to missing data, but had significantly higher SES (p=0.007, Cohen’s d=0.28).

### 3.2 Cerebral hemodynamics among patients, controls, and CHD subgroups

Inter-rater reliability was good for 4 and excellent for 6 of the measured cerebral hemodynamic parameters (mean ICC-coefficient=0.916, range=0.838–0.981). Supplementary Table 2 shows statistical estimates of the hemodynamic parameters among patients with CHD, cCHD, aCHD, and healthy controls (including the post-hoc measured VA). Supplementary Figure 3 shows a correlation matrix of the different hemodynamic parameters. Flow curves across the cardiac cycle for each vessel are plotted for every participant and group in supplementary Figure 4.

The linear regression models comparing cerebral hemodynamic parameters between patients and controls showed that the right ICA BF was significantly lower in patients with CHD, whereas the PBV of the left ICA was higher in the CHD group. However, standardized regression coefficients (β) revealed only small effect sizes (0.251 and 0.297), and after FDR-correction, there were no significant differences between the two groups (see Table 4 for statistical estimates). Further, post-hoc analyses revealed no significant interaction between group and sex (see supplementary Table 3).

**Table 4.**
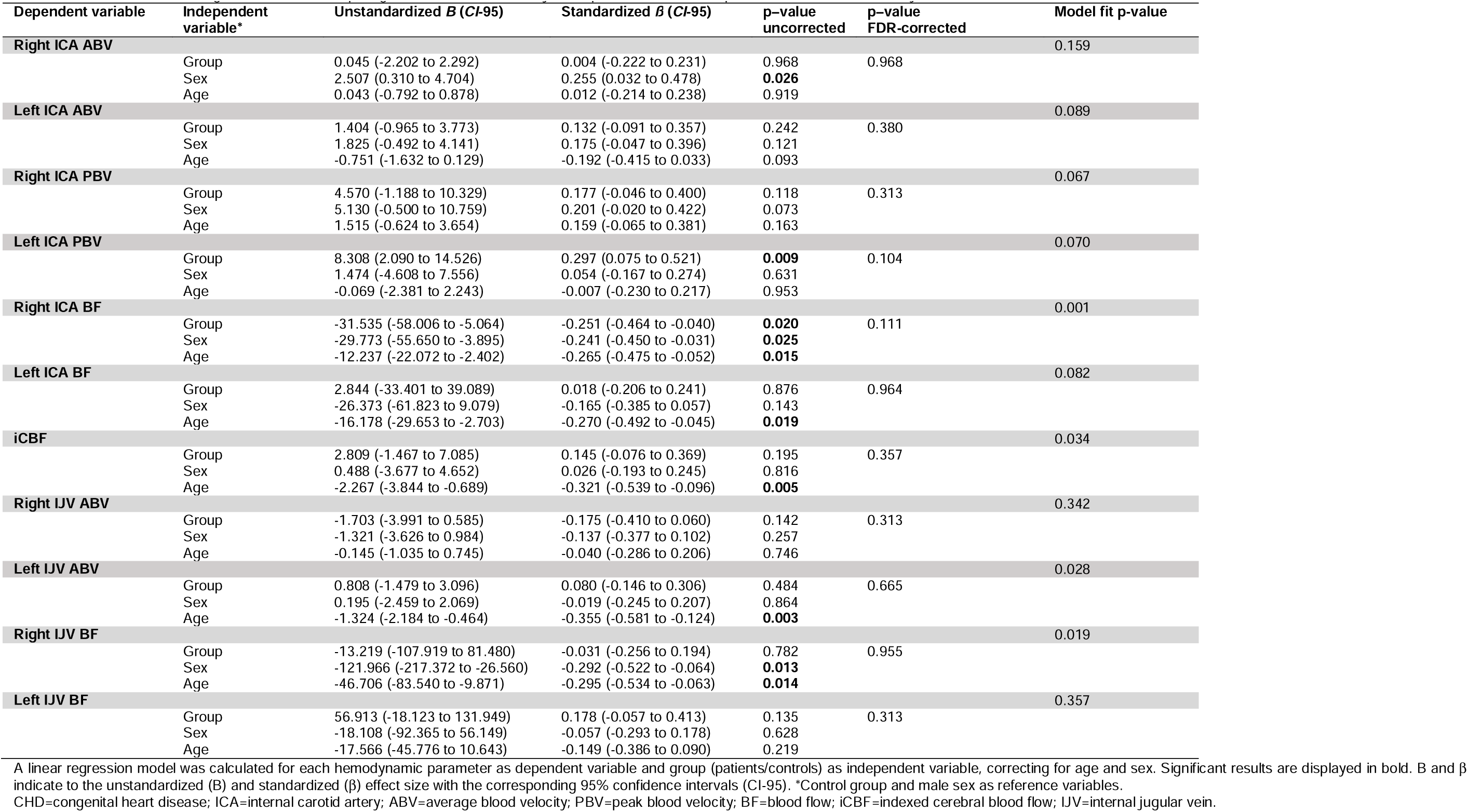
Results of the linear regression models comparing anterior cerebral hemodynamic parameters between patients with CHD and healthy controls.

The linear regression models comparing cerebral hemodynamic parameters between patients with cCHD, patients with aCHD and healthy controls showed that the right ICA BF was significantly lower in patients with cCHD compared to healthy controls, and left ICA PBV was significantly higher in patients with cCHD compared to healthy controls. ACHD had significantly higher left ICA BF, iCBF, left IJV ABV and left IJV BF compared to both cCHD and healthy controls. Effect sizes (β) were small to moderate (0.291–0.433) and after FDR-correction, there were no significant differences between any of the subgroups (see supplementary Table 4 for statistical estimates). Figure 2 and supplementary Figure 5 show forest plots with the standardized effects of cerebral hemodynamic parameters between patients with CHD and healthy controls (Figure 2) and between patients with aCHD, cCHD and healthy control (supplementary Figure 5). Figure 3 displays boxplots of those anterior cerebral hemodynamic parameters, which significantly differed between patients with CHD and healthy controls before correcting for multiple comparisons. Supplementary Figures 6 and 7 show boxplots of all the anterior hemodynamic parameters included in this study among groups or subgroups.

**Figure 2.**
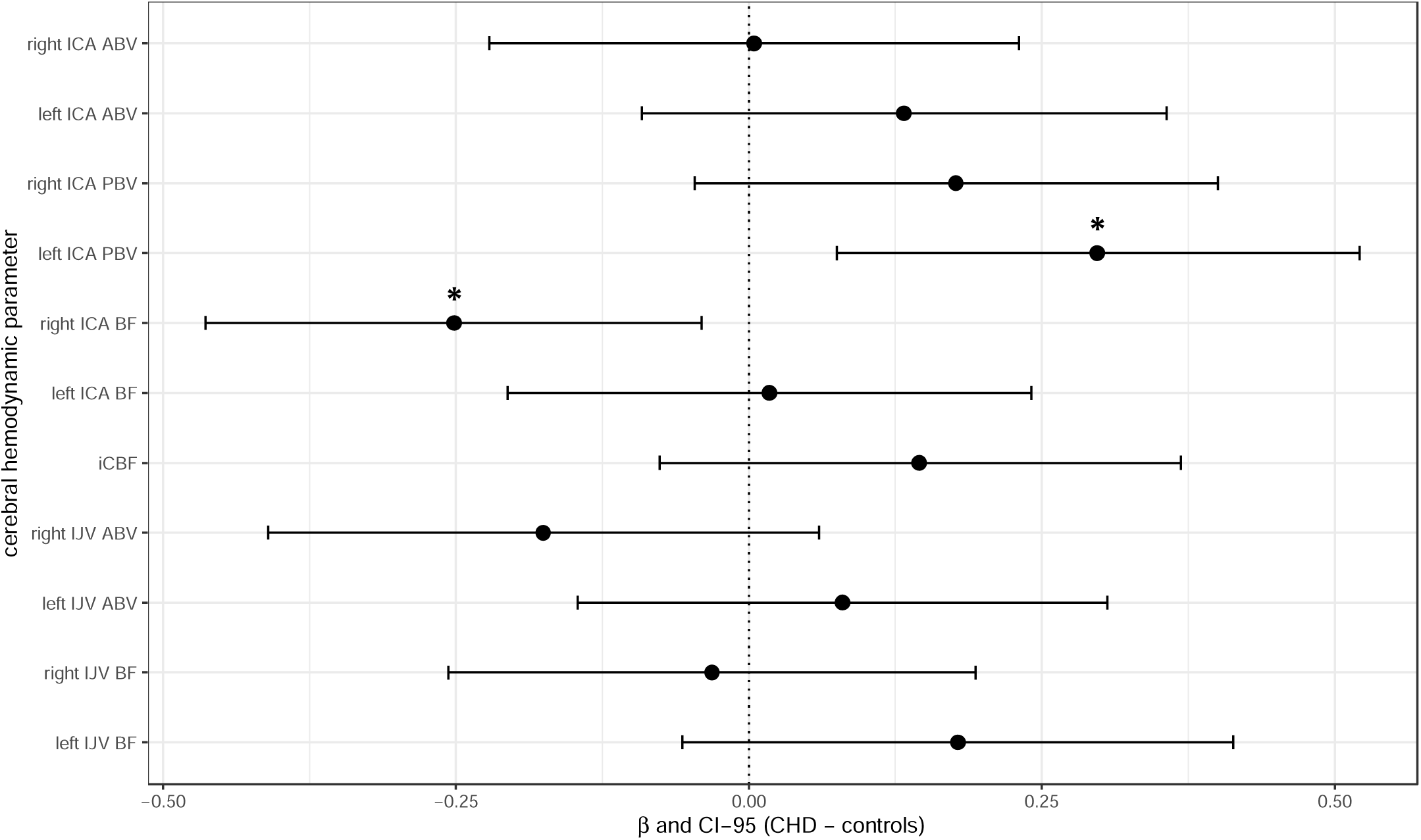
Forest plot of group effects in anterior cerebral hemodynamics. Forest plot displaying the standardized effect sizes (β=dot) and corresponding 95% confidence intervals (CI-95=lines) of the group effect when comparing anterior cerebral hemodynamic parameters of patients with CHD to healthy controls. Negative βs correspond to lower values for CHD, positive βs correspond to lower values for controls. Significant differences (without FDR-correction) are displayed with a *. Note that all effects were nonsignificant after FDR-correction. CHD=congenital heart disease; ICA=internal carotid artery; ABV=average blood velocity; PBV=peak blood velocity; BF=blood flow; iCBF=indexed cerebral blood flow; IJV=internal jugular vein.

**Figure 3.**
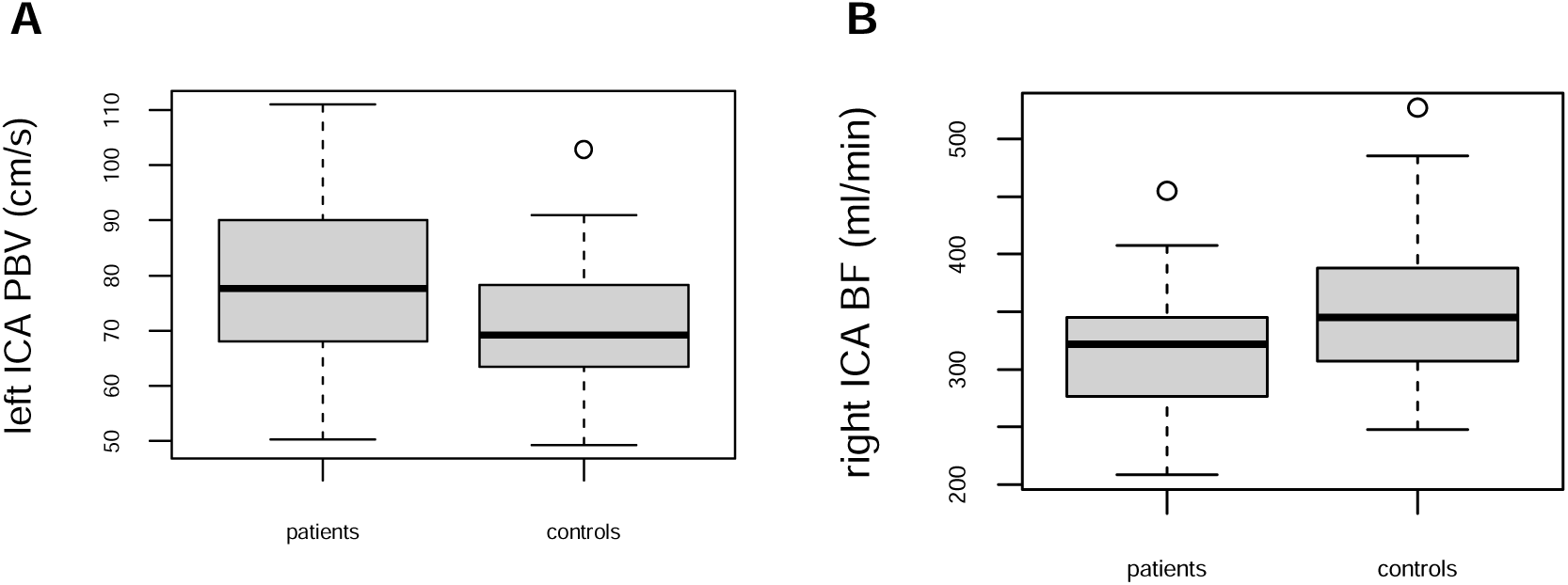
Boxplots of anterior cerebral hemodynamics. Boxplots displaying anterior cerebral hemodynamic parameters among groups that (without FDR-correction) significantly differed between patients with CHD and healthy controls. Note that all effects were nonsignificant after FDR-correction. CHD=congenital heart disease; ICA=internal carotid artery; PBV=peak blood velocity; BF=blood flow.

After accounting for the vessel angle, the post-hoc linear regression models assessing velocities and flow of the VA revealed significantly lower ABV and BF in the left VA in patients with CHD compared to controls (see Figure 4 for boxplots and see supplementary Table 5 for statistical estimates). These results remained significant after FDR-correction for both ABV and BF (β=0.248, respectively 0.314; FDR-corrected p-value=0.044, respectively 0.044).

**Figure 4.**
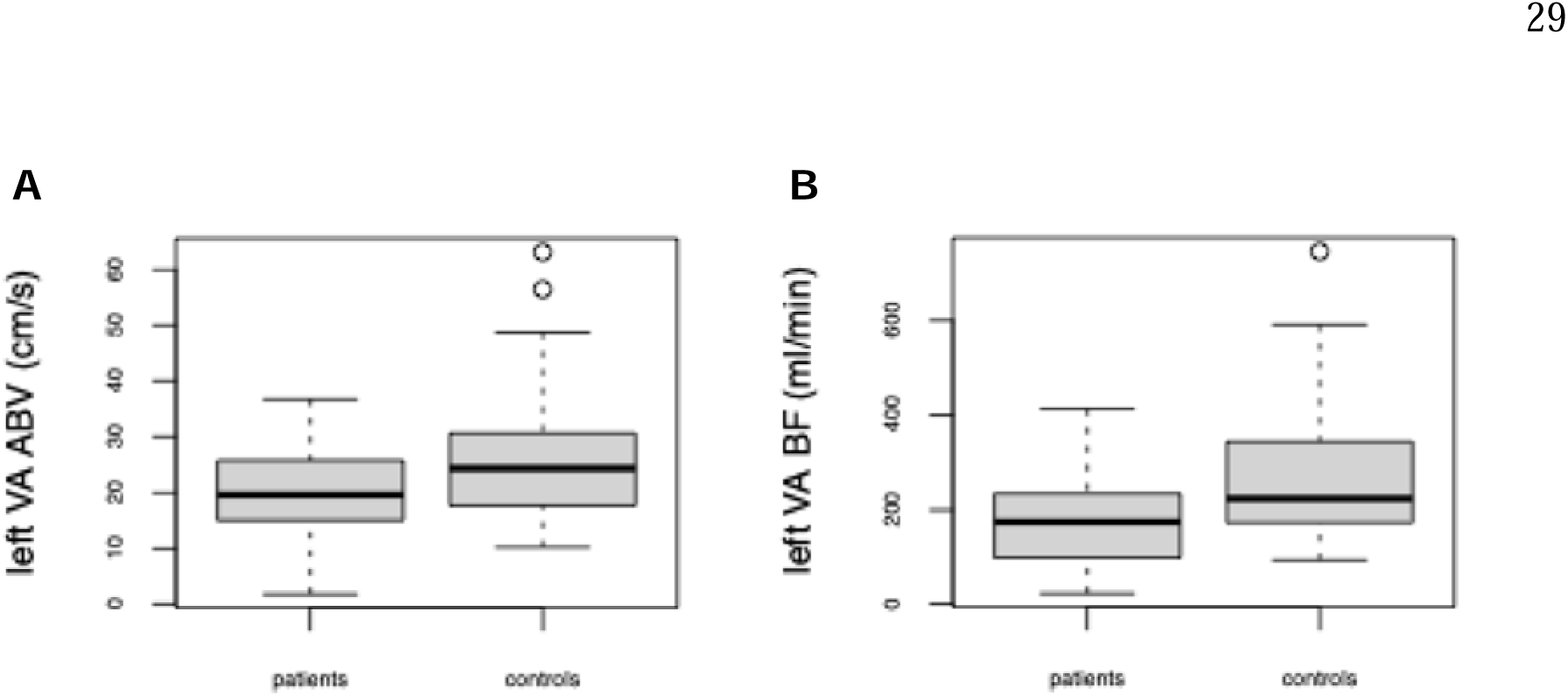
Boxplots of posterior cerebral hemodynamics. Boxplots displaying vertebral hemodynamic parameters among groups that significantly differed between patients with CHD and healthy controls. These effects were significant after FDR-correction (both corrected p-values=0.044). CHD=congenital heart disease; VA=vertebral artery; ABV=average blood velocity; BF=blood flow.

### 3.3 No relationship between cerebral hemodynamics and executive functions

Right ICA BF, left ICA BF, left ICA PBV, left ICA BF, iCBF, left IJV ABV and left IJV BF were included in regression models assessing the relationship between anterior cerebral hemodynamic parameters and executive functions as these measures displayed significant group or subgroup differences before FDR-correction. There was no significant correlation between any of the hemodynamic parameters and executive functions (see statistical estimates in supplementary Table 6).

## 4. Discussion

This study aimed to explore cerebral hemodynamics in adolescents with CHD who underwent open-heart surgery during their first year of life and wanted to investigate associations with executive function performance. In our cohort, after FDR-correction, there were no significant differences in anterior cerebral hemodynamic parameters between patients with CHD and healthy controls, or between patients with cCHD, patients with aCHD, and healthy controls. Before correcting for multiple comparisons, the overall CHD group showed lower BF of the right ICA and higher PBV of the left ICA than healthy controls, and there were multiple differences in hemodynamic factors between patients with aCHD, cCHD and healthy controls. All the differences in cerebral hemodynamic parameters were of small or moderate effect size. None of these hemodynamic parameters were significantly associated with the executive function summary score.

Alterations of cerebral hemodynamics have been shown preoperatively in infants with CHD, especially in the most severe cases of CHD involving a systemic-to-pulmonary shunt, such as those with cyanotic or univentricular CHD (De Silvestro et al., 2023). A recent study also confirmed that the presence of a systemic-to-pulmonary shunt is associated with reduced cerebral perfusion, by an amount dependent on the size of the shunt. In contrast, the presence of an aortic arch obstruction and altered oxygen saturation were not associated with altered perfusion (De Silvestro et al., 2024). Two recent studies have suggested that reduced cerebral perfusion persists in adolescence and early adulthood (Easson et al., 2023; Schmithorst et al., 2022). Of note, both studies used ASL to assess cerebral hemodynamics, in comparison to pcMRI used in our study. Easson and colleagues reported moderate to large effect sizes in their regional analyses with ASL. In contrast, using pcMRI we only identified a few group differences between patients and controls of small effect sizes by using pcMRI. These group effects did not survive FDR-correction. A possible explanation for preserved anterior cerebral hemodynamics at this age could be the stable medical condition of adolescents that had infant open-heart surgery during childhood. When looking at the distribution of cardiac complications in patients with CHD, incident rates are generally low in later childhood and adolescence and increase with older age. Some complications, including stroke, show a U-formed curve, with a decrease of prevalence after early childhood and an increase after adolescence (Arslani et al., 2018). In fact, within our sample only one patient showed a valve insufficiency and one showed decreased cardiac function in the echocardiographic examination at the time of assessment. This compensated cardiologic state, compared to infants or adults with CHD, may provide sufficient blood supply to the brain. Importantly, our study only included patients of a younger age (up to 15 years) than the two studies finding reduced cerebral perfusion in adolescents and young adults with CHD (up to 21 and 24 years) (Easson et al., 2023; Schmithorst et al., 2022). Schmithorst et al. (2022) reported a steeper age-related decline of perfusion during late adolescence in patients with CHD compared to healthy controls. Since it is well known that cerebral perfusion decreases with age (Satterthwaite et al., 2014; Tarumi and Zhang, 2018), along with the rising risk for cardiac complications (Arslani et al., 2018), this might result in impaired cerebral hemodynamics in patients with CHD only at a later age than assessed in our sample. We further did not find a significant interaction effect of the gender when comparing patients with CHD to healthy controls, such as described by Easson et al. (2023), who found reduced cerebral perfusion in female but not in male patients with CHD. Sex specific differences in cerebral hemodynamics have been found to evolve during mid-puberty in the healthy population (Satterthwaite et al., 2014), and therefore sex specific alterations of cerebral hemodynamics might only appear at a later stage of adolescence in patients with CHD. Of note, our sample mostly included patients who had corrective cardiac surgery and only five cases with univentricular CHD who had palliative cardiac surgery. Easson et al. (2023) and Schmithorst et al. (2022) both found cerebral perfusion to be lower in univentricular compared to biventricular patients with CHD, due to reasons such as elevated central venous pressure and subsequent altered systemic venous flow in patients with a Fontan circulation. We did not compare univentricular to biventricular patients due to the small sample of individuals with univentricular CHD (n=5). Cerebral hemodynamics have been shown to change with cardiac surgery in patients with CHD (Cheng et al., 2020; Sun et al., 2022; Yavasoglu and Can, 2021). According to one study, cerebral blood velocities return to normal values in patients with ventricular or atrial septal defect and significantly improve in patients with Tetralogy of Fallot 3–6 months after corrective heart surgery (Yavasoglu and Can, 2021). Our results suggest that anterior cerebral hemodynamics remain preserved or are not strongly altered during early adolescence and after corrective open-heart surgery.

Although we did not find any significant differences in anterior cerebral hemodynamics between patients with CHD, subgroups of CHD and healthy controls after correcting for multiple comparisons, we still found some differences with small to moderate effect sizes (standardized β-values 0.251–0.433) before FDR-correction. In combination with the limited sample size, FDR-correction might have been too conservative for detecting these effects. Therefore, the patients with CHD assessed in our study might still have slight to moderate alterations of anterior cerebral hemodynamics. This could indicate that, despite changes after cardiac surgery (Cheng et al., 2020; Sun et al., 2022; Yavasoglu and Can, 2021), blood supply to the brain might still be slightly impaired in adolescents with CHD, especially in patients with cCHD, who presented with lower arterial and venous ABV and BF in several vessels compared to patients with aCHD or controls. This could contribute to the elevated risk of cerebrovascular accidents, which has been described for patients with CHD even beyond childhood and is most prominent in patients with cyanotic CHD (Hoffmann et al., 2010). In contrast, arterial PBV was increased in patients with CHD (relative to controls) and cCHD (relative to controls) in our sample. Elevated PBV could possibly indicate the presence of central arterial stiffness, which has been found in vessels such as the carotid arteries in patients with CHD (Sandhu et al., 2021). Within our sample, patients with aCHD showed higher values of ABV and BF in several vessels as well as higher iCBF than healthy controls. We did not find other studies among any age group describing similar findings, and do not have a plausible explanation for this phenomenon. Therefore, these results may be random, or due to chance, likely attributed to the relatively small number of patients with aCHD (n=12) included in this study. Importantly, however, none of those differences were statistically significant after correcting for multiple comparisons and are therefore hypothesis generating for future studies rather than evidence providing.

We further did not find any evidence that cerebral hemodynamics correlate with executive functions. Our results suggest that cerebral hemodynamics are not among the risk factors leading to impaired executive functions in adolescents aged 10–15 years with CHD. Possibly, other factors, such as hypoxemia, play a more important role for neurodevelopmental outcome among young adolescents with CHD (Wernovsky, 2006).

Our post-hoc analysis of the vertebral flow indicated that the CHD group may have lower left VA ABV and BF than controls. However, this result should be considered with caution until it can be replicated in an independent sample, with optimal slice positioning for the VA. Nevertheless, the apparent reduction in the posterior cerebral circulation in combination with preserved anterior hemodynamics in patients with CHD is interesting, since subclavian steal phenomenon with reduced or inversed unilateral vertebral flow has been described in infants with CHD such as coarctation of the aorta and interruption of the aortic arch (Deeg et al., 1993). Preserved anterior circulation with alterations in left-posterior flow could reflect a residuum of early alterations in vertebral hemodynamics in patients with CHD, especially in those with an underlying aortic pathology. However, as mentioned above, those considerations are speculative, but could provide hypotheses for further studies.

Overall, our results suggest that anterior cerebral hemodynamics are not seriously impaired in early adolescents with CHD who underwent infant open-heart surgery, whereas the posterior cerebral circulation might be altered. Also, alterations of cerebral hemodynamics do not seem to be among the driving reasons for impaired executive functions in adolescents with CHD. There is a need for future studies with bigger sample sizes to detect potential mild alterations of cerebral hemodynamics in adolescents with CHD and their clinical relevance. Future studies should also assess broader age ranges and analyze cerebral hemodynamics longitudinally and in correlation to cardiac and vascular function, in order to further model the age-related trajectory of hemodynamic alterations and to evaluate the predictive value of cerebral hemodynamics for neurodevelopmental outcomes.

## 5. Limitations

A major limitation of our study is the restricted statistical power achieved with our modest sample size in relation to the small to moderate effect sizes of our analyses. Also, the small number of univentricular cases prevented us from examining cerebral hemodynamics in univentricular CHD after palliative heart surgery, and the narrow age range of our participants precluded the analysis of cerebral hemodynamics in an older population. The control participants were slightly younger than the patients. However, all analyses were controlled for age and sex. Further, we were only able to measure the blood velocities and flow in the VA post-hoc after correcting for the vessel angle, which may have led to a less precise quantification compared to images with optimal slice positioning. Due to safety requirements, we were not able to perform an ASL sequence for patients with certain cardiac implants. This prevented us from analyzing regional cerebral perfusion and its association with domain-specific cognitive performance, which have been stated to correlate in children and adolescents with CHD (Schmithorst et al., 2022). Finally, cerebral hemodynamics and executive function performance were assessed concurrently and thus, we could not infer about any long-term, predictive value of pcMRI for neurodevelopmental outcome.

## 6. Conclusions

This study suggests that there are no or only mild alterations of anterior cerebral hemodynamics in adolescents with CHD who underwent infant open-heart surgery, whereas the posterior cerebral perfusion is reduced in early adolescents with CHD. Further we did not find evidence of an association between cerebral hemodynamics and executive function performance in these patients. Similar cerebral hemodynamics between patients and healthy peers could be related to the stable cardiac condition of adolescents that underwent corrective infant heart surgery. Well-powered, longitudinal trials are needed to investigate age-related trajectory of hemodynamic alterations across the life span and to further evaluate the predictive value of cerebral hemodynamics for neurodevelopmental outcomes.

## Data Availability

Data cannot be made publicly available due to patient privacy and ethical regulations. De-identified data and analysis scripts will be shared with qualified researchers upon reasonable request and subject to ethical approval.

## CRediT Author Contributions

François Mojon: Conceptualization, Formal analysis, Investigation, Data curation, Writing – Original Draft, Visualization

Beatrice Latal, MD MPH: Conzeptualization, Methology, Resources, Writing – Review & Editing, Funding acquisition

Raimund Kottke, MD: Investigation, Validation, Writing – Review & Editing

Oliver Kretschmar, MD: Conzeptualization, Validation, Writing – Review & Editing

Ruth Tuura O’Gorman, PhD: Conzeptualization, Methology, Validation, Resources, Writing – Review & Editing, Supervision, Funding acquisition

Melanie Ehrler, PhD: Conzeptualization, Methology, Validation, Formal analysis, Writing – Review & Editing, Visualization, Supervision, Project administration

## Acknowledgments

We thank all the participating adolescents and their parents.

## Abbreviations

ABV: average blood velocity;
aCHD: acyanotic congenital heart disease;
ASL: arterial spin labeling;
BF: blood flow;
cCHD: cyanotic congenital heart disease;
CHD: congenital heart disease;
FDR: False Discovery Rate;
ICA: internal carotid artery;
iCBF: indexed cerebral blood flow;
ICC: intraclass correlation;
IJV: internal jugular vein;
PBV: peak blood velocity;
pcMRI: phase-contrast magnetic resonance imaging;
VA: vertebral artery.

