## Supplementary Material for "Cerebral hemodynamics in early adolescents with congenital heart disease after infant open-heart surgery"

**Supplementary Tables**

**Supplementary Table 1.** Neuropsychological test battery to assess executive functions performance.

| **Executive function domains** | **Neuropsychological test** | **Test measurement** |
| --- | --- | --- |
| Working memory | Digit Span forward & backward (WISC-IV)  Letter-number sequencing (WISC-IV)  Corsi Block Tapping Test (Corsi) | Number of correct items  Number of correct items  Number of correct items |
| Inhibition | Subtest Interference, Colour Word Interference Task (D-KEFS) Go/NoGo (TAP) | Completion time  Number of commission errors |
| Cognitive flexibility | Subtest Letter-Number-Switching of the Trail Making Task (D-KEFS)  TAP Flexibility (TAP) | Completion time  Median reaction time |
| Fluency | Subtests s-words and animals (RWT)  Subtest filled-dots-only, Design Fluency Test (D-KEFS) | Number of correct items  Number of correct items |
| Planning | Tower Task (D-KEFS) | Total achievement score |

WISC-IV=Wechsler Intelligence Scale for Children Fourth Edition (Petermann and Petermann, 2008), Corsi (1972), D-KEFS=Delis-Kaplan Executive Function System (Delis et al., 2001), TAP=Test of Attentional Performance (Zimmermann and Fimm, 1994), RWT=Regensburger verbal fluency Test (Aschenbrenner et al., 2000).

**Supplementary Table 2.** Mean values, SD, and number of missing values of different hemodynamic parameters among patients with CHD, cCHD, aCHD, and healthy controls.

| Hemodynamic parameter | Total CHD (n=34) | Cyanotic CHD (n=22) | Acyanotic CHD (n=12) | Controls (n=49) |
| --- | --- | --- | --- | --- |
| Right ICA ABV, cm/s, mean (SD), [missing, n] | 23.15 (5.03), [1] | 23.05 (5.54), [1] | 23.32 (4.20), [0] | 23.44 (4.89), [0] |
| Left ICA ABV, cm/s, mean (SD), [missing, n] | 25.10 (5.93), [1] | 24.62 (6.31), [1] | 25.96 (5.34), [0] | 24.43 (4.76), [0] |
| Right ICA PBV, cm/s, mean (SD), [missing, n] | 72.19 (13.75), [1] | 72.47 (13.30), [1] | 71.68 (15.08), [0] | 67.41 (11.82), [0] |
| Left ICA PBV, cm/s, mean (SD), [missing, n] | 78.84 (15.54), [1] | 81.63 (16.24), [1] | 73.97 (13.49), [0] | 70.79 (11.54), [0] |
| Right ICA BF, ml/min, mean (SD), [missing, n] | 317.01 (56.19), [1] | 308.89 (49.63), [1] | 331.20 (66.05), [0] | 351.87 (62.14), [0] |
| Left ICA BF, ml/min, mean (SD), [missing, n] | 346.85 (94.46), [1] | 324.40 (85.37), [1] | 386.12 (100.31), [0] | 350.05 (70.02), [0] |
| iCBF, ml/min/100g, mean (SD), [missing, n] | 57.36 (10.74), [2] | 54.82 (8.76), [2] | 61.60 (12.68), [0] | 56.02 (8.68), [0] |
| Right IJV ABV, cm/s, mean (SD), [missing, n] | 10.31 (3.87), [2] | 9.91 (3.39), [0] | 11.17 (4.86), [2] | 11.91 (5.32), [4] |
| Left IJV ABV, cm/s, mean (SD), [missing, n] | 11.25 (3.79), [2] | 9.94 (2.16), [2] | 13.44 (4.90), [0] | 11.34 (5.75), [3] |
| Right IJV BF, ml/min, mean (SD), [missing, n] | 395.11 (219.65), [2] | 401.07 (206.33), [0] | 382.00 (258.02), [2] | 420.69 (201.68), [4] |
| Left IJV BF, ml/min, mean (SD), [missing, n] | 327.33 (149.73), [2] | 285.31 (124.27), [2] | 397.37 (167.17), [0] | 280.21 (162.15), [3] |
| Right VA ABV, cm/s, mean (SD), [missing, n] | 23.624 (9.366), [1] | 23.804 (8.527), [0] | 23.265 (11.304), [1] | 25.788 (10.437), [1] |
| Left VA ABV, cm/s, mean (SD), [missing, n] | 20.167 (8.096), [2] | 21.275 (7.761), [1] | 18.052 (8.672), [1] | 26.335 (10.707), [2] |
| Right VA PBV, cm/s, mean (SD), [missing, n] | 85.064 (46.879), [1] | 83.327 (40.022), [0] | 88.539 (60.405), [1] | 87.789 (91.492), [1] |
| Left VA PBV, cm/s, mean (SD), [missing, n] | 81.404 (42.722), [2] | 87.483 (35.23), [1] | 69.798 (54.312), [1] | 92.362 (48.626), [2] |
| Right VA BF, ml/min, mean (SD), [missing, n] | 235.612 (114.155), [1] | 215.102 (114.155), [0] | 276.631 (165.229), [1] | 267.068 (191.044), [1] |
| Left VA BF, ml/min, mean (SD), [missing, n] | 179.075 (94.238), [2] | 177.847 (79.561), [1] | 181.421 (121.907), [1] | 272.45 (138.016), [2] |

CHD=congenital heart disease; cCHD=cyanotic congenital heart disease; aCHD=acyanotic congenital heart disease; ICA=internal carotid artery; ABV=average blood velocity; PBV=peak blood velocity; BF=blood flow; iCBF=indexed cerebral blood flow; IJV=internal jugular vein; VA=vertebral artery.

**Supplementary Table 3.** Results of the linear regression models assessing the interaction between group and sex on cerebral hemodynamic parameters.

| **Dependent variable** | **Independent**  **variable** | **B (CI-95)** | **ß (CI-95)** | **p-value**  **uncorrected** | **regression model p-value** |
| --- | --- | --- | --- | --- | --- |
| **Right ICA ABV** | Group:sex | 2.220 (-2.252 to 6.691) | 0.161 (-0.163 to 0.484) | 0.326 | 0.190 |
| **Left ICA ABV** | Group:sex | 0.055 (-4.692 to 4.801) | 0.004 (-0.319 to 0.326) | 0.982 | 0.167 |
| **Right ICA PBV** | Group:sex | 10.074 (-1.227 to 21.375) | 0.281 (-0.034 to 0.596) | 0.080 | 0.037 |
| **Left ICA PBV** | Group:sex | 5.756 (-6.635 to 18.147) | 0.148 (-0.171 to 0.468) | 0.358 | 0.097 |
| **Right ICA BF** | Group:sex | 5.384 (-47.606 to 58.373) | 0.031 (-0.274 to 0.335) | 0.840 | 0.004 |
| **Left ICA BF** | Group:sex | -39.645 (-111.715 to 32.424) | -0.176 (-0.495 to 0.144) | 0.277 | 0.096 |
| **iCBF** | Group:sex | -3.677 (-12.186 to 4.832) | -0.138 (-0.458 to 0.182) | 0.392 | 0.053 |
| **Right IJV ABV** | Group:sex | 1.394 (-3.199 to 5.987) | 0.102 (-0.234 to 0.438) | 0.547 | 0.450 |
| **Left IJV ABV** | Group:sex | -3.447 (-7.971 to 1.077) | -0.241 (-0.558 to 0.075) | 0.133 | 0.023 |
| **Right IJV BF** | Group:sex | -80.361 (-270.006 to 109.283) | -0.136 (-0.457 to 0.185) | 0.401 | 0.032 |
| **Left IJV BF** | Group:sex | -121.578 (-269.607 to 26.450) | -0.270 (-0.598 to 0.059) | 0.106 | 0.209 |

Post-hoc, a linear regression model was calculated for each anterior hemodynamic parameter as dependent variable and a group:sex interaction term (group=patients/controls) as independent variable, correcting for age (not displayed here). B and β indicate to the unstandardized (B) and standardized (β) effect size with the corresponding 95% confidence intervals (CI-95).

ICA=internal carotid artery; ABV=average blood velocity; PBV=peak blood velocity; BF=blood flow; iCBF=indexed cerebral blood flow; IJV=internal jugular vein.

**Supplementary Table 4.** Results of the linear regression models comparing anterior cerebral hemodynamic parameters between patients with cCHD, aCHD and healthy controls.

| **Dependent variable** | **Independent**  **variable*** | **Unstandardized *B* (CI-95)** | **Standardized *ß* (CI-95)** | **p-value**  **uncorrected** | **p-value**  **FDR-corrected** | **Model fit p-value** |
| --- | --- | --- | --- | --- | --- | --- |
| **Right ICA ABV** |  |  |  |  |  | 0.273 |
|  | CCHD – ACHD | 0.040 (-3.516 to 3.596) | 0.003 (-0.254 to 0.260) | 0.982 | 0.982 |  |
|  | CCHD – controls | -0.031 (-2.612 to 2.551) | -0.003 (-0.262 to 0.256) | 0.981 | 0.982 |  |
|  | ACHD – controls | -0.071 (-3.306 to 3.164) | -0.007 (-0.332 to 0.317) | 0.965 | 0.820 |  |
| **Left ICA ABV** |  |  |  |  |  | 0.124 |
|  | CCHD – ACHD | 1.652 (-2.085 to 5.388) | 0.112 (-0.142 to 0.366) | 0.382 | 0.742 |  |
|  | CCHD – controls | -0.827 (-3.535 to 1.882) | -0.078 (-0.333 to 0.177) | 0.545 | 0.787 |  |
|  | ACHD – controls | -2.478 (-5.875 to 0.918) | -0.234 (-0.554 to 0.087) | 0.150 | 0.330 |  |
| **Right ICA PBV** |  |  |  |  |  | 0.120 |
|  | CCHD – ACHD | -2.080 (-11.180 to 7.019) | -0.058 (-0.311 to 0.196) | 0.650 | 0.858 |  |
|  | CCHD – controls | -5.298 (-11.904 to 1.308) | -0.205 (-0.460 to 0.051) | 0.114 | 0.290 |  |
|  | ACHD – controls | -3.218 (-11.495 to 5.060) | -0.124 (-0.444 to 0.196) | 0.441 | 0.766 |  |
| **Left ICA PBV** |  |  |  |  |  | 0.047 |
|  | CCHD – ACHD | -7.935 (-17.628 to 1.758) | -0.205 (-0.455 to 0.045) | 0.107 | 0.290 |  |
|  | CCHD – controls | -11.081 (-18.106 to -4.056) | -0.397 (-0.648 to -0.145) | **0.002** | 0.068 |  |
|  | ACHD – controls | -3.146 (-11.957 to 5.664) | -0.113 (-0.428 to 0.203) | 0.479 | 0.787 |  |
| **Right ICA BF** |  |  |  |  |  | 0.001 |
|  | CCHD – ACHD | 32.072 (-9.177 to 73.321) | 0.184 (-0.053 to 0.421) | 0.126 | 0.296 |  |
|  | CCHD – controls | 42.754 (12.806 to 72.701) | 0.341 (0.102 to 0.58) | **0.006** | 0.068 |  |
|  | ACHD – controls | 10.682 (-26.841 to 48.205) | 0.085 (-0.214 to 0.384) | 0.572 | 0.787 |  |
| **Left ICA BF** |  |  |  |  |  | 0.008 |
|  | CCHD – ACHD | 74.784 (19.878 to 129.689) | 0.332 (0.088 to 0.575) | **0.008** | 0.068 |  |
|  | CCHD – controls | 23.294 (-16.499 to 63.088) | 0.143 (-0.101 to 0.388) | 0.247 | 0.510 |  |
|  | ACHD – controls | -51.489 (-101.396 to -1.582) | -0.317 (-0.624 to -0.010) | **0.043** | 0.143 |  |
| **iCBF** |  |  |  |  |  | 0.005 |
|  | CCHD – ACHD | 8.201 (1.685 to 14.717) | 0.308 (0.063 to 0.553) | **0.014** | 0.068 |  |
|  | CCHD – controls | 0.142 (-4.614 to 4.897) | 0.007 (-0.239 to 0.253) | 0.953 | 0.982 |  |
|  | ACHD – controls | -8.06 (-13.935 to -2.184) | -0.417 (-0.721 to -0.113) | **0.008** | 0.068 |  |
| **Right IJV ABV** |  |  |  |  |  | 0.416 |
|  | CCHD – ACHD | 1.434 (-2.247 to 5.116) | 0.101 (-0.158 to 0.360) | 0.440 | 0.766 |  |
|  | CCHD – controls | 2.139 (-0.415 to 4.692) | 0.220 (-0.043 to 0.484) | 0.099 | 0.290 |  |
|  | ACHD – controls | 0.704 (-2.736 to 4.144) | 0.073 (-0.282 to 0.427) | 0.684 | 0.869 |  |
| **Left IJV ABV** |  |  |  |  |  | 0.004 |
|  | CCHD – ACHD | 4.321 (0.899 to 7.744) | 0.313 (0.065 to 0.561) | **0.014** | 0.068 |  |
|  | CCHD – controls | 0.744 (-1.785 to 3.272) | 0.074 (-0.176 to 0.323) | 0.560 | 0.787 |  |
|  | ACHD – controls | -3.577 (-6.691 to -0.464) | -0.354 (-0.661 to -0.046) | **0.025** | 0.091 |  |
| **Right IJV BF** |  |  |  |  |  | 0.043 |
|  | CCHD – ACHD | 7.124 (-145.888 to 160.137) | 0.012 (-0.237 to 0.260) | 0.926 | 0.982 |  |
|  | CCHD – controls | 15.384 (-90.720 to 121.489) | 0.037 (-0.216 to 0.289) | 0.773 | 0.945 |  |
|  | ACHD – controls | 8.260 (-134.709 to 151.229) | 0.020 (-0.321 to 0.360) | 0.909 | 0.982 |  |
| **Left IJV BF** |  |  |  |  |  | 0.004 |
|  | CCHD – ACHD | 126.684 (13.436 to 239.931) | 0.291 (0.031 to 0.552) | **0.014** | 0.068 |  |
|  | CCHD – controls | -11.408 (-95.079 to 72.263) | -0.036 (-0.298 to 0.227) | 0.560 | 0.787 |  |
|  | ACHD – controls | -138.092 (-241.108 to -35.075) | -0.433 (-0.756 to -0.110) | **0.025** | 0.091 |  |

A linear regression model was calculated for each hemodynamic parameter as dependent variable and subgroup (cCHD/aCHD/controls) as independent variable, correcting for age and sex (not displayed here). Significant results are displayed in bold. B and β indicate to the unstandardized (B) and standardized (β) effect size with the corresponding 95% confidence intervals (CI-95). *In CCHD – ACHD, CCHD – controls and ACHD – controls, the first named subgroup is the reference variable.

cCHD=cyanotic congenital heart disease; aCHD=acyanotic congenital heart disease; ICA=internal carotid artery; ABV=average blood velocity; PBV=peak blood velocity; BF=blood flow; iCBF=indexed cerebral blood flow; IJV=internal jugular vein.

**Supplementary Table 5.** Results of the linear regression models comparing vertebral hemodynamic parameters between patients with CHD and healthy controls.

| **Dependent variable** | **Independent**  **variable*** | **Unstandardized *B* (*CI*-95)** | **Standardized *ß* (*CI*-95)** | **p–value**  **uncorrected** | **p–value**  **FDR-corrected** | **Model fit p-value** |
| --- | --- | --- | --- | --- | --- | --- |
| **Right VA ABV** |  |  |  |  |  | 0.111 |
|  | Group | -0.784 (-5.364 to 3.796) | -0.039 (-0.265 to 0.188) | 0.734 | 0.303 |  |
|  | Sex | 2.041 (-2.451 to 6.534) | 0.102 (-0.122 to 0.326) | 0.368 |  |  |
|  | Age | -1.636 (-3.352 to 0.079) | -0.219 (-0.446 to 0.011) | 0.061 |  |  |
| **Left VA ABV** |  |  |  |  |  | 0.003 |
|  | Group | -5.102 (-9.554 to -0.650) | -0.248 (-0.466 to -0.032) | **0.025** | **0.044** |  |
|  | Sex | 5.191 (0.830 to 9.551) | 0.256 (0.041 to 0.471) | **0.020** |  |  |
|  | Age | -0.683 (-2.336 to 0.970) | -0.091 (-0.307 to 0.127) | 0.413 |  |  |
| **Right VA PBV** |  |  |  |  |  | 0.725 |
|  | Group | -1.683 (17.068 to 34.226) | -0.011 (-0.244 to 0.222) | 0.926 | 0.880 |  |
|  | Sex | -17.383 (-52.605 to 17.839) | -0.114 (-0.345 to 0.117) | 0.329 |  |  |
|  | Age | -5.069 (-18.519 to 8.381) | -0.089 (-0.324 to 0.147) | 0.455 |  |  |
| **Left VA PBV** |  |  |  |  |  | 0.275 |
|  | Group | -7.652 (-29.433 to 14.129) | -0.082 (-0.314 to 0.151) | 0.486 | 0.424 |  |
|  | Sex | 15.808 (-5.524 to 37.140) | 0.170(-0.060 to 0.401) | 0.144 |  |  |
|  | Age | -2.169 (-10.255 to 5.918) | -0.063 (-0.295 to 0.170) | 0.595 |  |  |
| **Right VA BF** |  |  |  |  |  | 0.462 |
|  | Group | -17.994 (-97.464 to 61.475) | -0.052 (-0.284 to 0.179) | 0.653 | 0.633 |  |
|  | Sex | -2.981 (-80.930 to 4.967) | -0.009 (-0.238 to 0.221) | 0.939 |  |  |
|  | Age | -20.606 (-50.372 to 9.159) | -0.162 (-0.395 to 0.072) | 0.172 |  |  |
| **Left VA BF** |  |  |  |  |  | 0.005 |
|  | Group | -82.577 (-140.159 to -24.995) | -0.314 (-0.534 to -0.095) | **0.006** | **0.044** |  |
|  | Sex | 29.748 (-26.646 to 86.142) | 0.114 (-0.103 to 0.331) | 0.297 |  |  |
|  | Age | -10.706 (-32.084 to 10.673) | -0.112 (-0.329 to 0.109) | 0.322 |  |  |

Post-hoc, a linear regression model was calculated for each vertebral hemodynamic parameter as dependent variable and group (patients/controls) as independent variable, correcting for age and sex. Significant results are displayed in bold. B and β indicate to the unstandardized (B) and standardized (β) effect size with the corresponding 95% confidence intervals (CI-95). *Control group and male sex as reference variables.

CHD=congenital heart disease; VA=vertebral artery; ABV=average blood velocity; PBV=peak blood velocity; BF=blood flow.

**Supplementary Table 6.** Results of the linear regression models assessing the relationship between cerebral hemodynamic parameters and executive functions.

| **Independent**  **variable** | **B (CI-95)** | **ß (CI-95)** | **p-value**  **uncorrected** | **p-value**  **FDR-corrected** | **regression model p-value** |
| --- | --- | --- | --- | --- | --- |
| **Left ICA PBV** | 0.010 (-0.007 to 0.028) | 0.119 (-0.081 to 0.311) | 0.245 | 0.815 | <0.001 |
| **Right ICA BF** | 0.001 (-0.004 to 0.005) | 0.033 (-0.183 to 0.249) | 0.761 | 0.815 | <0.001 |
| **Left ICA BF** | 0.001 (-0.002 to 0.004) | 0.061 (-0.141 to 0.259) | 0.557 | 0.815 | <0.001 |
| **iCBF** | -0.013 (-0.039 to 0.014) | -0.098 (-0.297 to 0.105) | 0.346 | 0.815 | <0.001 |
| **Left IJV ABV** | 0.016 (-0.035 to 0.067) | 0.063 (-0.142 to 0.270) | 0.539 | 0.815 | <0.001 |
| **Left IJV BF** | 0.000 (-0.001 to 0.002) | 0.023 (-0.175 to 0.221) | 0.815 | 0.815 | <0.001 |

A linear regression model was calculated with executive functions as dependent variable and different anterior hemodynamic parameters as independent variables, correcting for age, sex, group (patients/controls), and SES (not displayed here).

B and β indicate to the unstandardized (B) and standardized (β) effect size with the corresponding 95% confidence intervals (CI-95).

ICA=internal carotid artery; ABV=average blood velocity; PBV=peak blood velocity; BF=blood flow; iCBF=indexed cerebral blood flow; IJV=internal jugular vein; SES=socio-economic status.

**Supplementary Figures**

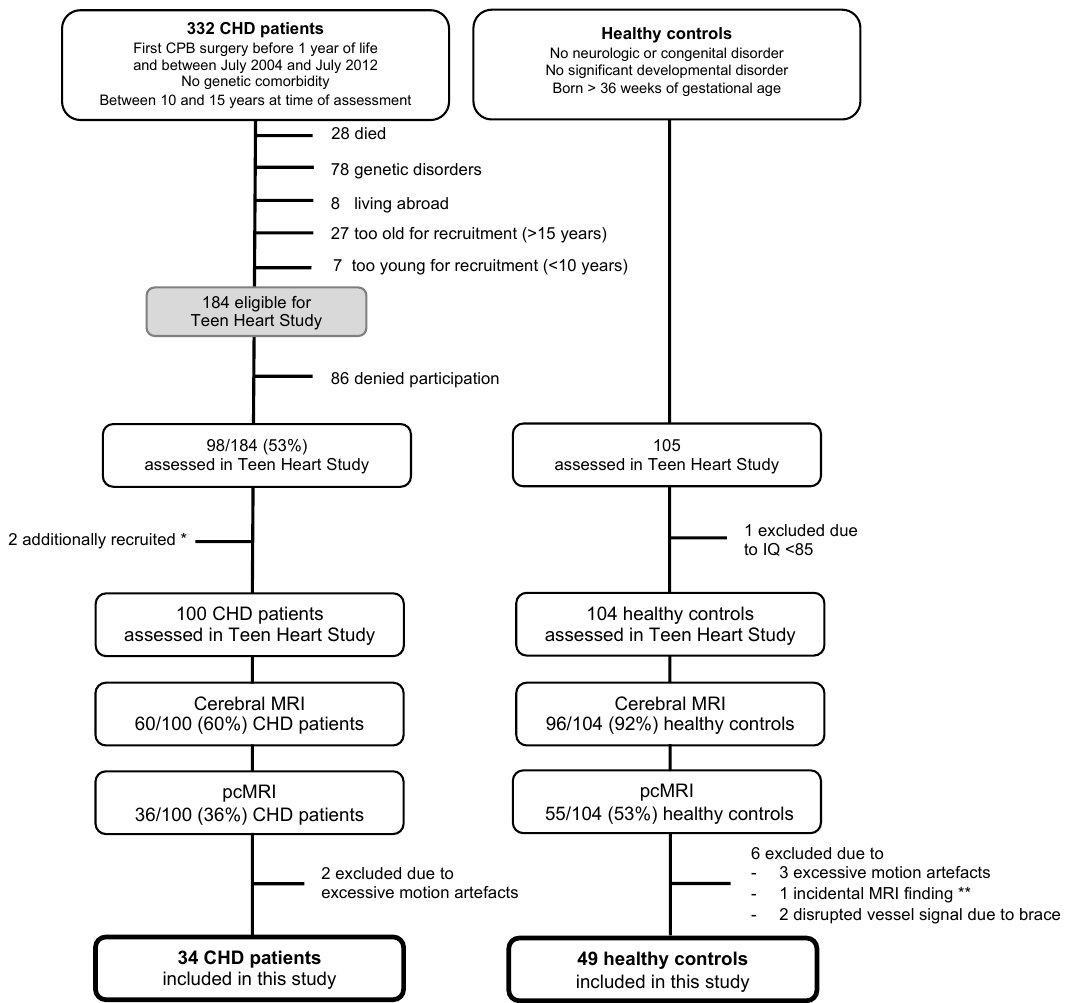

**Supplementary Figure 1. Recruitment procedure.**

Patients were recruited from two longitudinal cohort studies conducted at the University Children’s Hospital Zurich. a) Spillmann et al. (2021) b) von Rhein et al. (2020). * 2 patients were additionally recruited from the developmental outpatient clinic with the same inclusion criteria but families did not originally participate in the cohort study. ** Big cystic lesion in the posterior horn of the left lateral ventricle.

CHD=congenital heart disease; CPB=cardiopulmonary bypass; MRI=magnetic resonance imaging; pcMRI=phase-contrast magnetic resonance imaging.

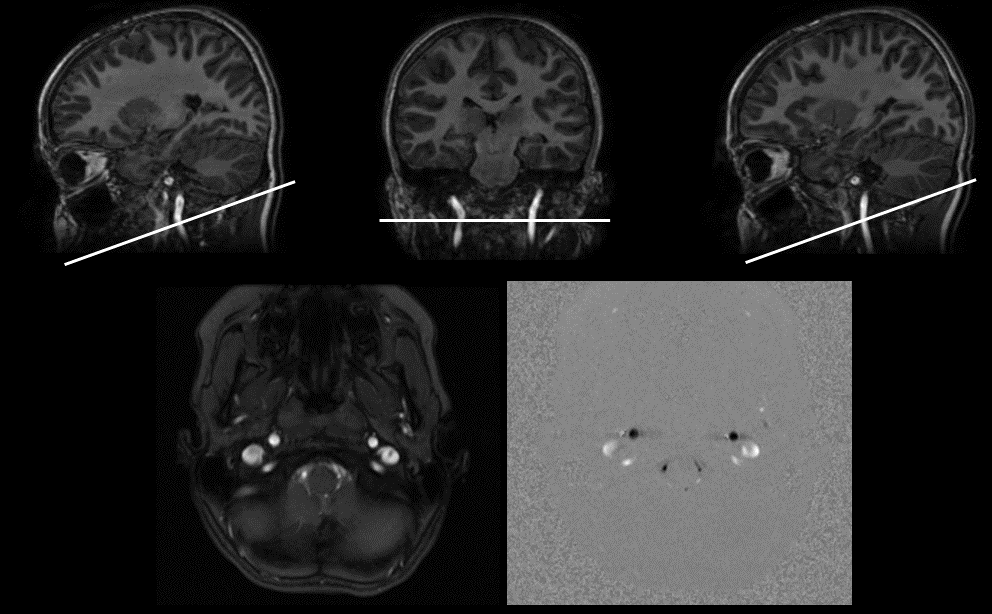

**Supplementary Figure 2. T1-weighted scan and acquisition of pcMRI.**

Axial phase-contrast images (bottom right) were acquired using a single-slice sequence near the base of the cerebellum, perpendicular to the internal carotid arteries and internal jugular veins, as visualized on a sagittal and coronal reformat of a three-dimensional T1-weighted scan.

pcMRI=phase-contrast magnetic resonance imaging.

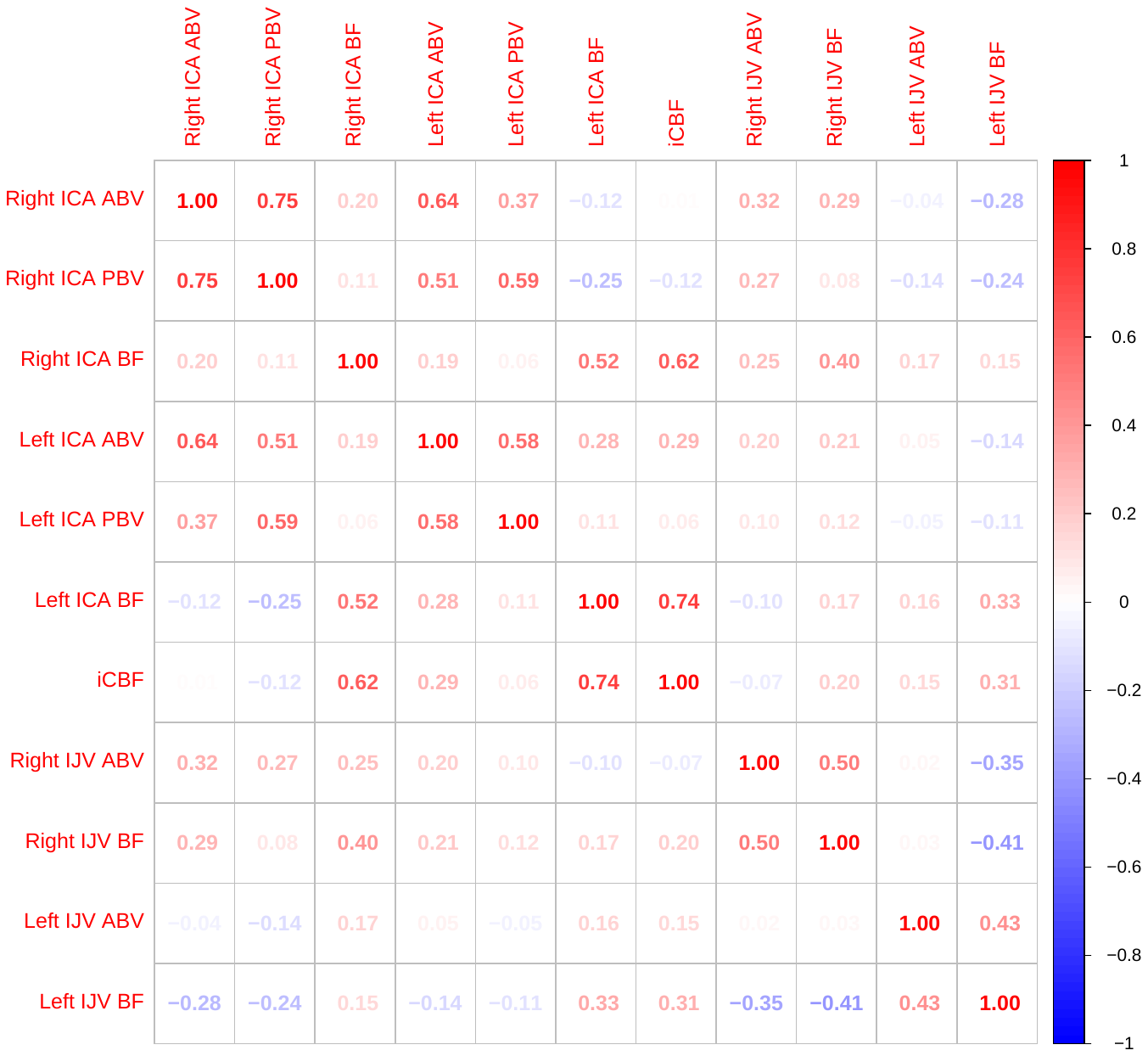

**Supplementary Figure 3. Correlation matrix of hemodynamic parameters.**
Correlation matrix with all the hemodynamic parameters primary assessed in this study.

ICA=internal carotid artery; ABV=average blood velocity; PBV=peak blood velocity; BF=blood flow; iCBF=indexed cerebral blood flow; IJV=internal jugular vein.

| 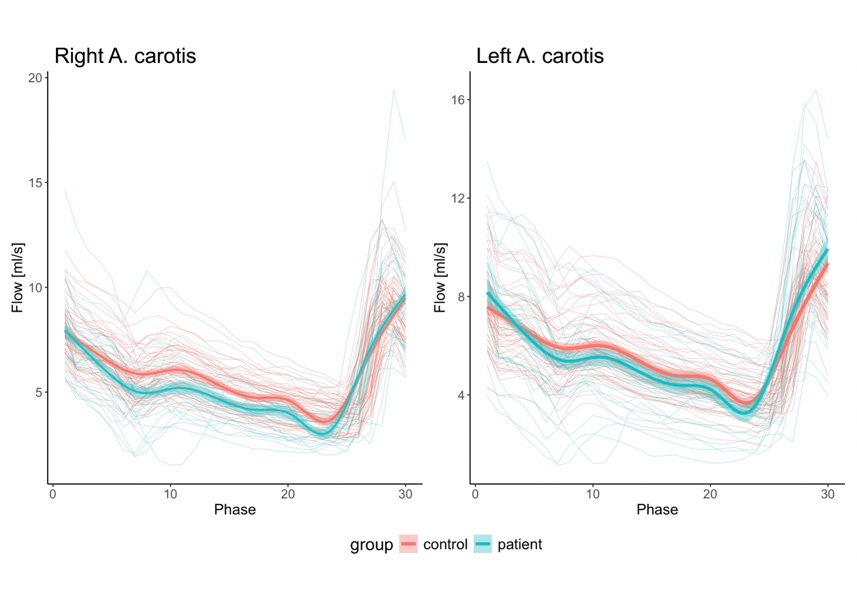**A** | 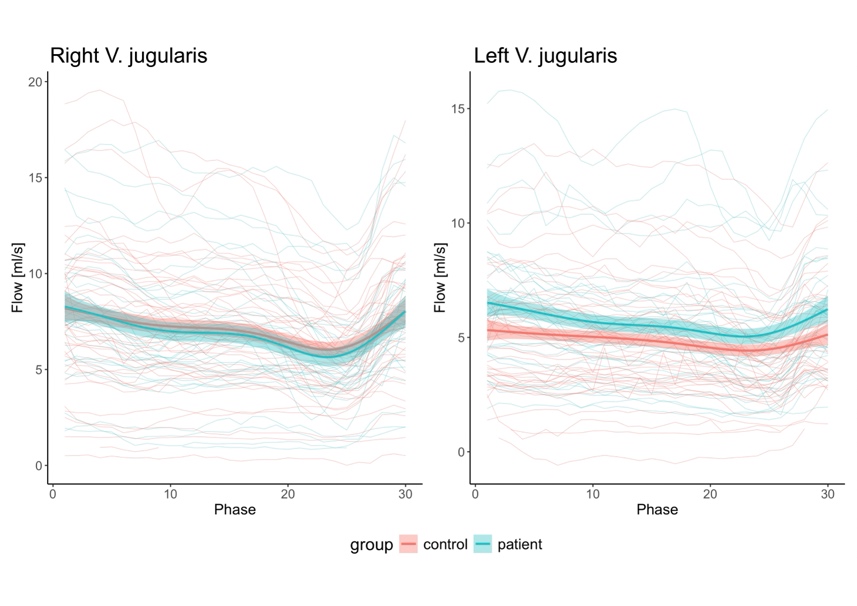**B** |
| --- | --- |
| 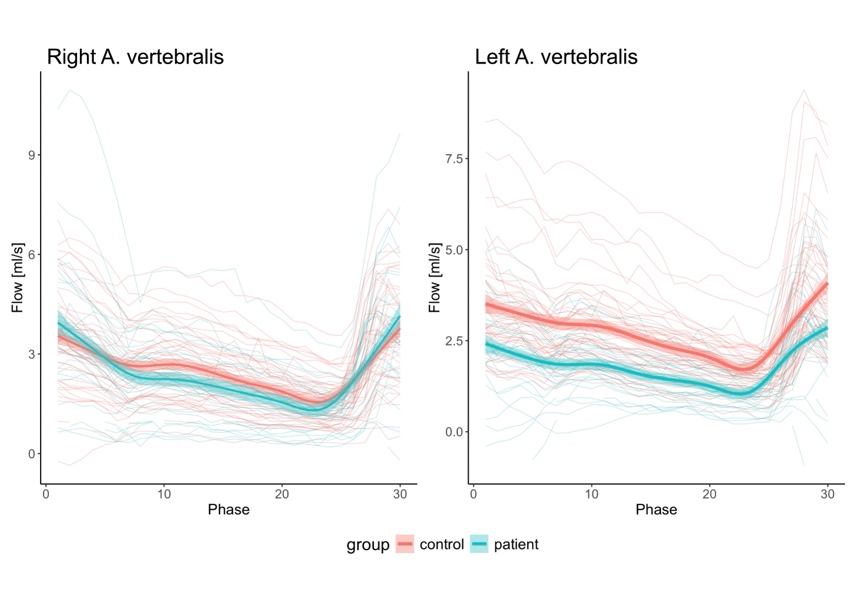**C** |  |

**Supplementary Figure 4. Flow curves across the cardiac cycle.**

Curves showing the blood flow across the cardiac cycle for each vessel for every participant and group (control = healthy controls, patient = patients with congenital heart disease).

| 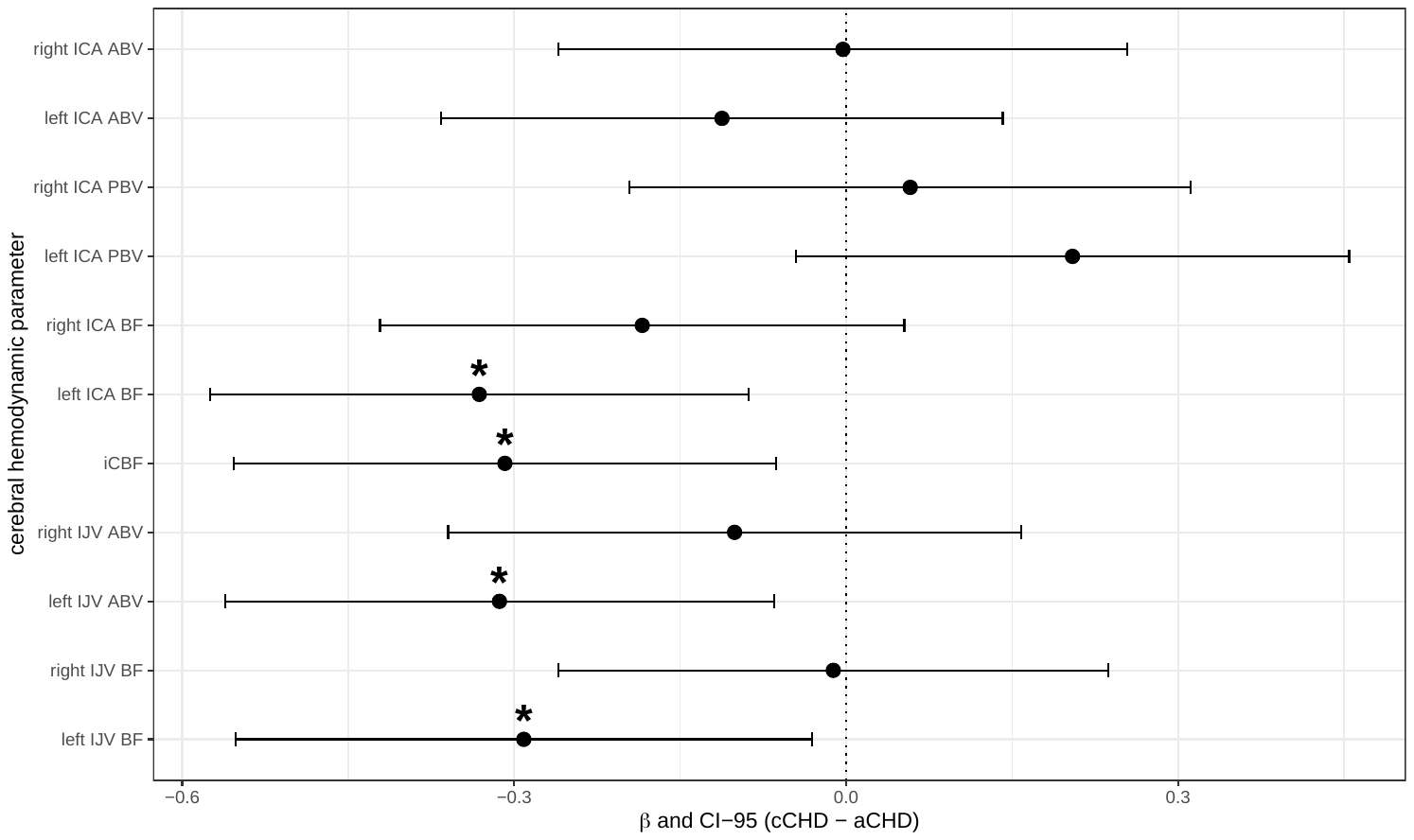**A** | 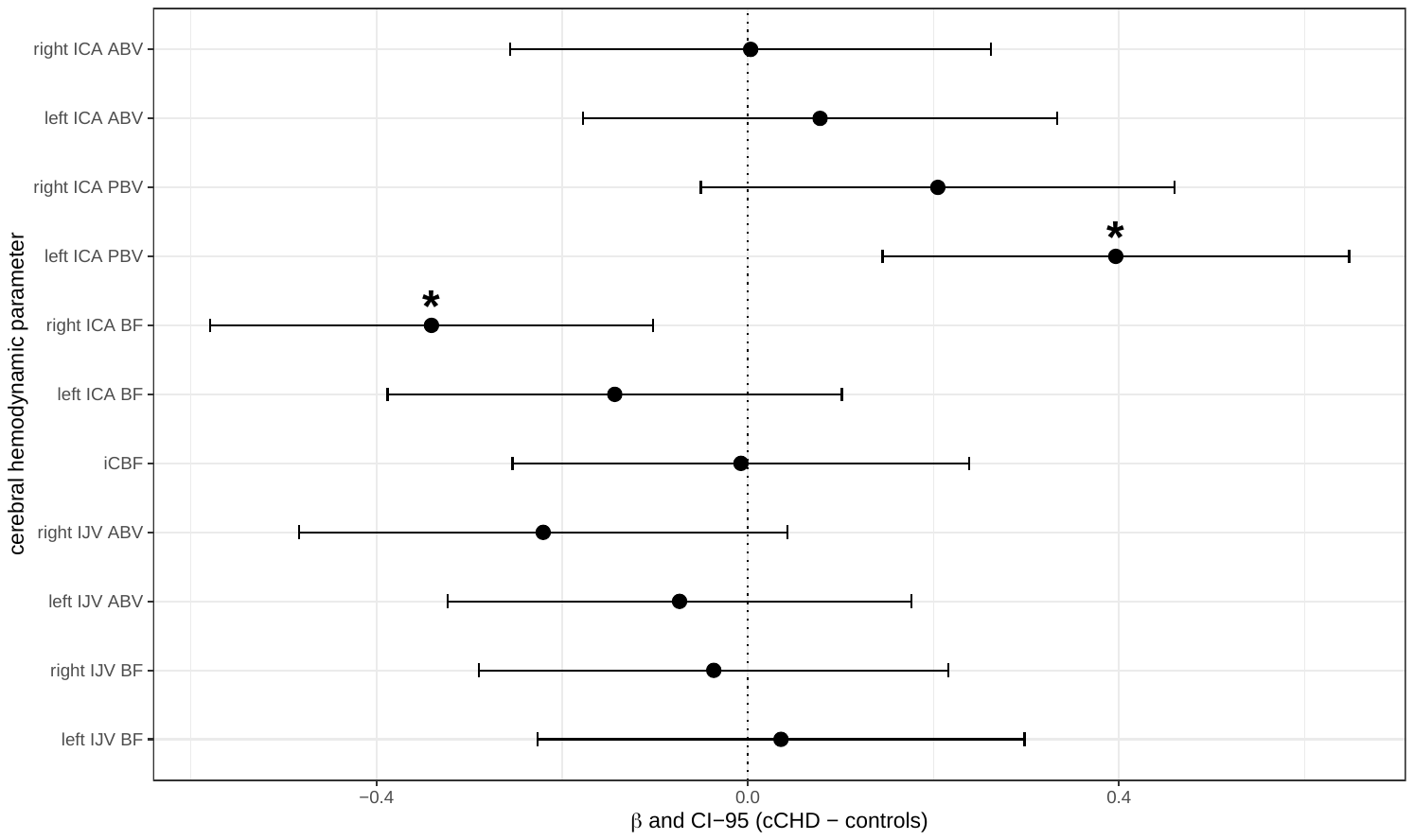**B** |
| --- | --- |
| 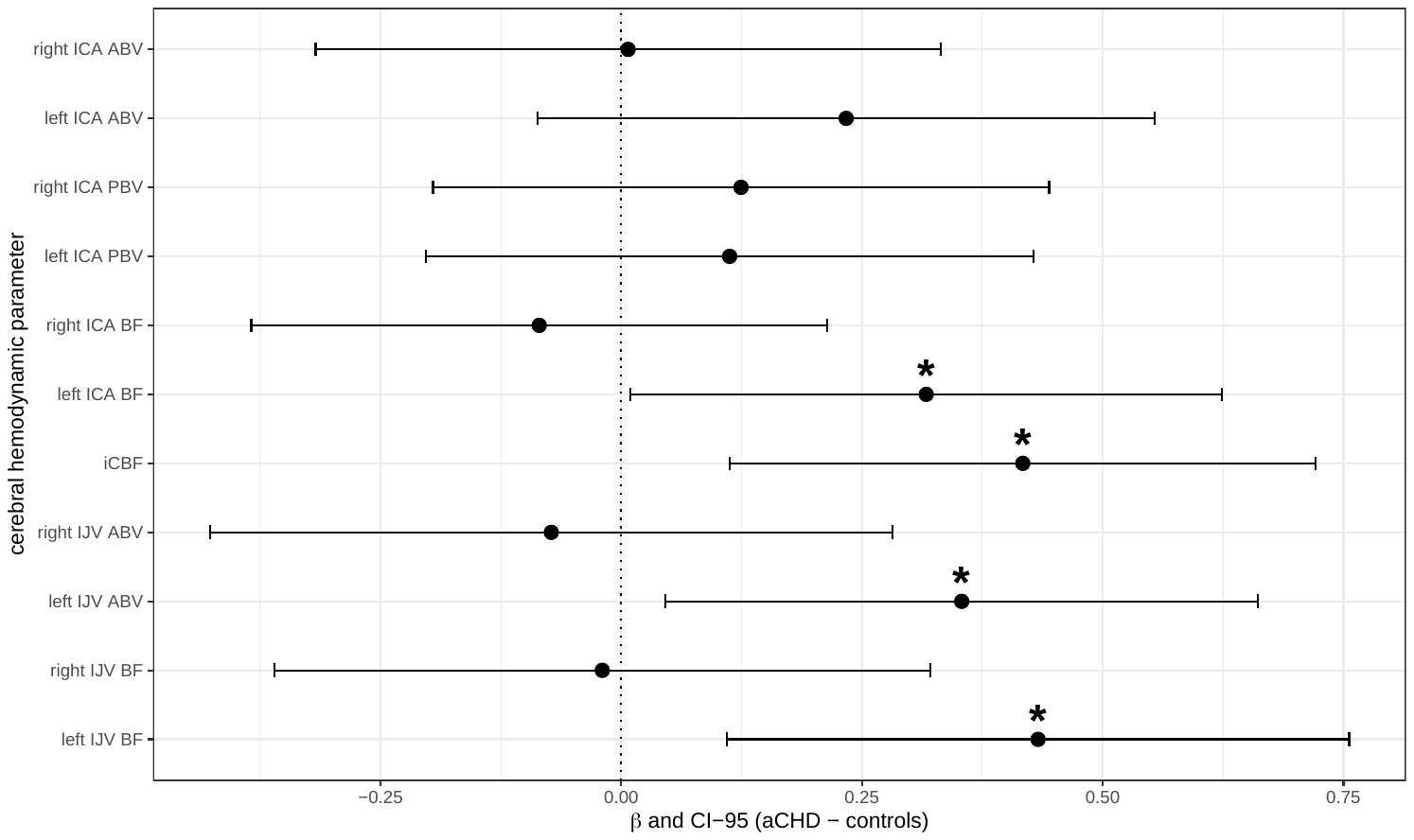**C** |  |

**Supplementary Figure 5. Forest plots of subgroup effects in anterior cerebral hemodynamics.**

Forests plots displaying the standardized effect sizes (β=dot) and corresponding 95% confidence intervals (CI-95=lines) of the subgroup effect when comparing different cerebral hemodynamic parameters of cCHD to aCHD (A), cCHD to healthy controls (B), or aCHD to healthy controls (C). Negative βs correspond to lower values for cCHD (A and B), or aCHD (C), positive βs correspond to lower values for aCHD (A) or controls (B and C). Significant differences (without FDR-correction) are displayed with a *****. Note that all effects were nonsignificant after FDR-correction.

cCHD=cyanotic congenital heart disease; aCHD=acyanotic congenital heart disease; ICA=internal carotid artery; ABV=average blood velocity; PBV=peak blood velocity; BF=blood flow; iCBF=indexed cerebral blood flow; IJV=internal jugular vein.

| 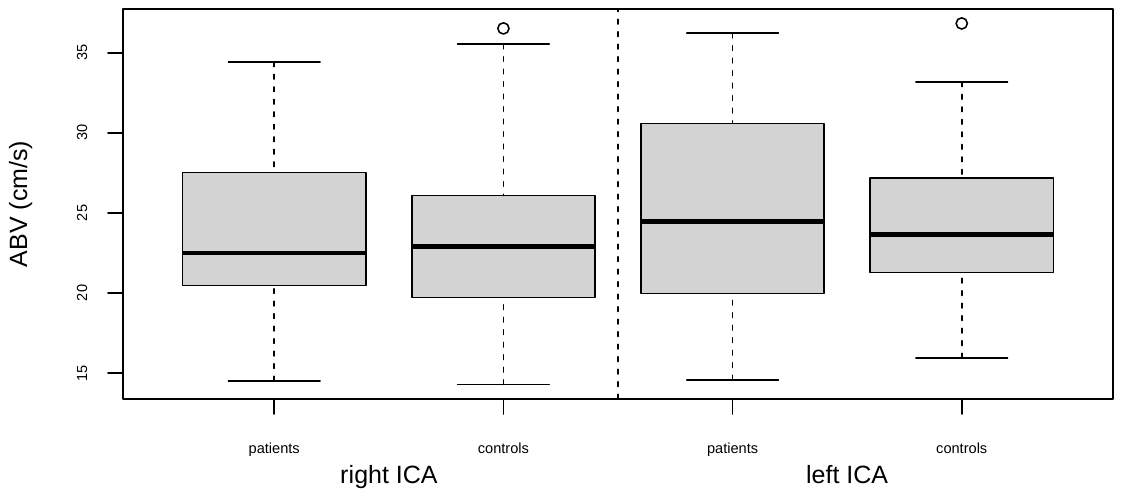**A** | 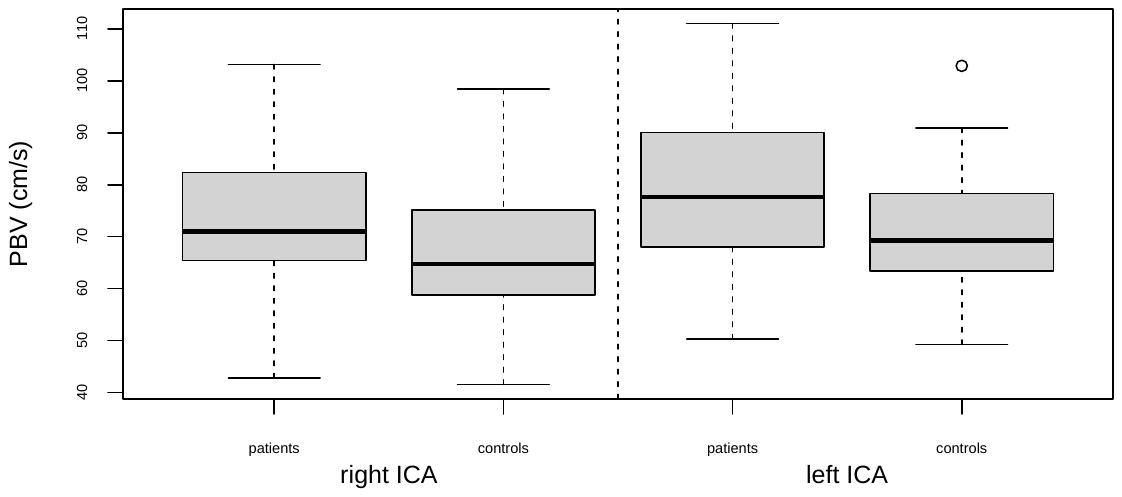**B** |
| --- | --- |
| 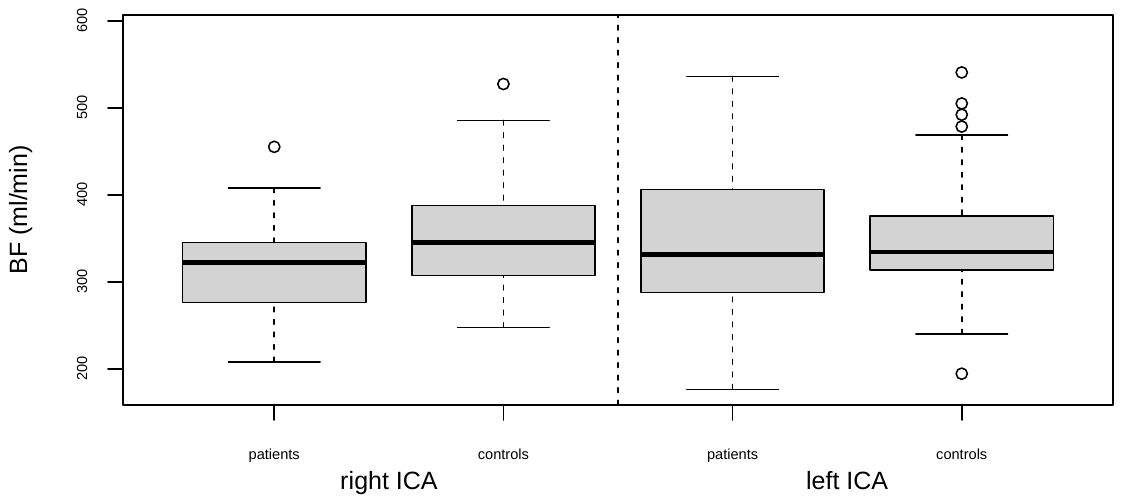**C** | 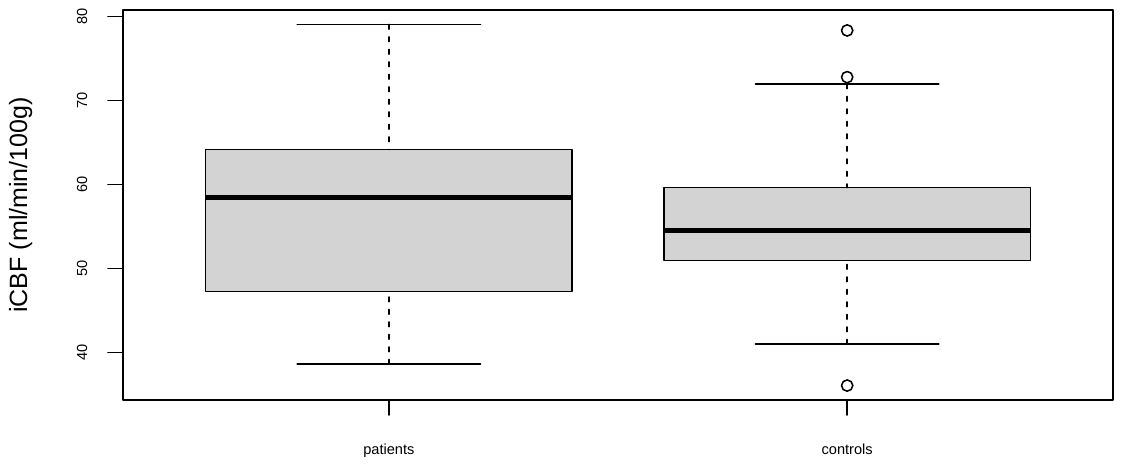**D** |
| 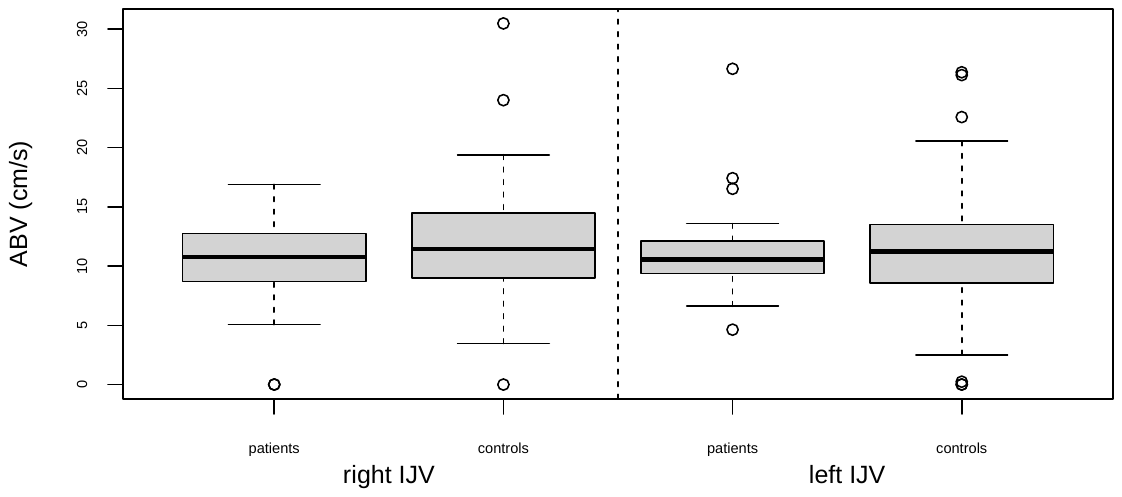**E** | 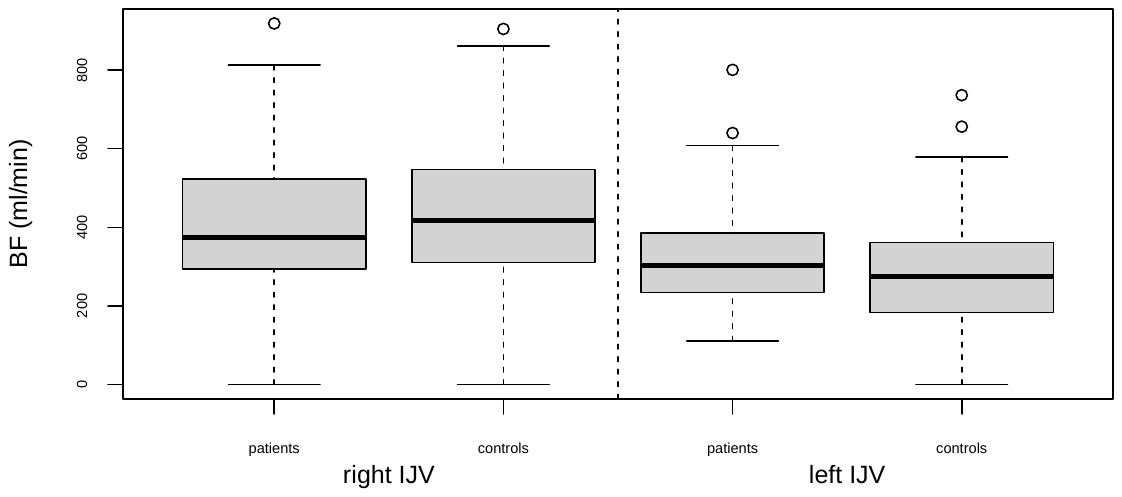**F** |

**Supplementary Figure 6. Boxplots of anterior cerebral hemodynamics among groups.**

Boxplots displaying all the cerebral hemodynamic parameters that were primary assessed in this study among patients with CHD and healthy controls.

CHD=congenital heart disease; ICA=internal carotid artery; ABV=average blood velocity; PBV=peak blood velocity; BF=blood flow; iCBF=indexed cerebral blood flow; IJV=internal jugular vein.

| 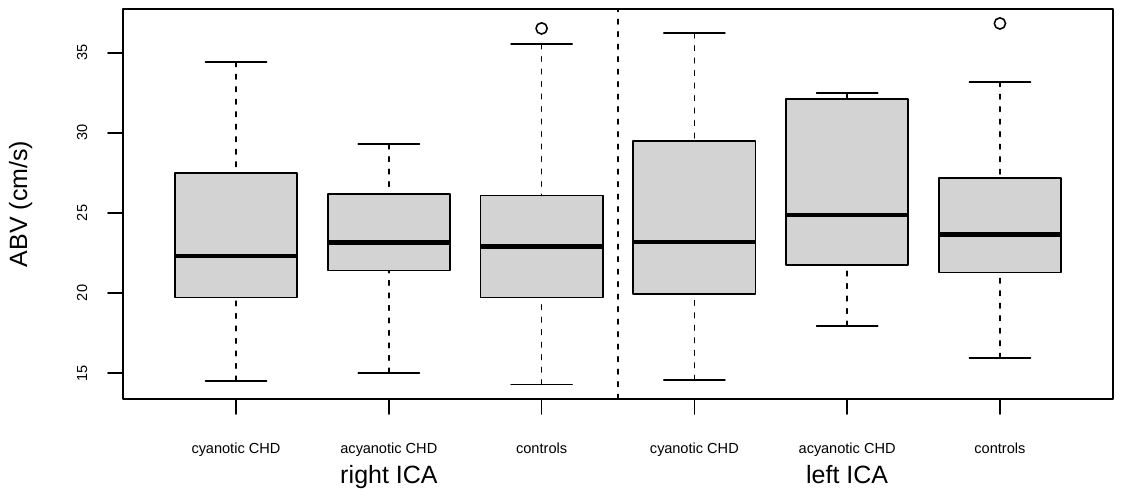**A** | 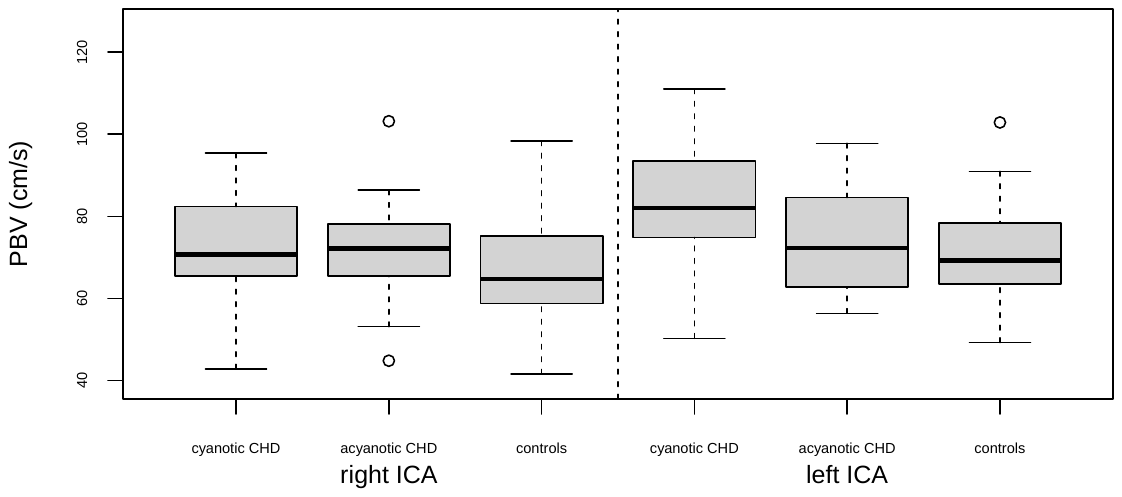**B** |
| --- | --- |
| 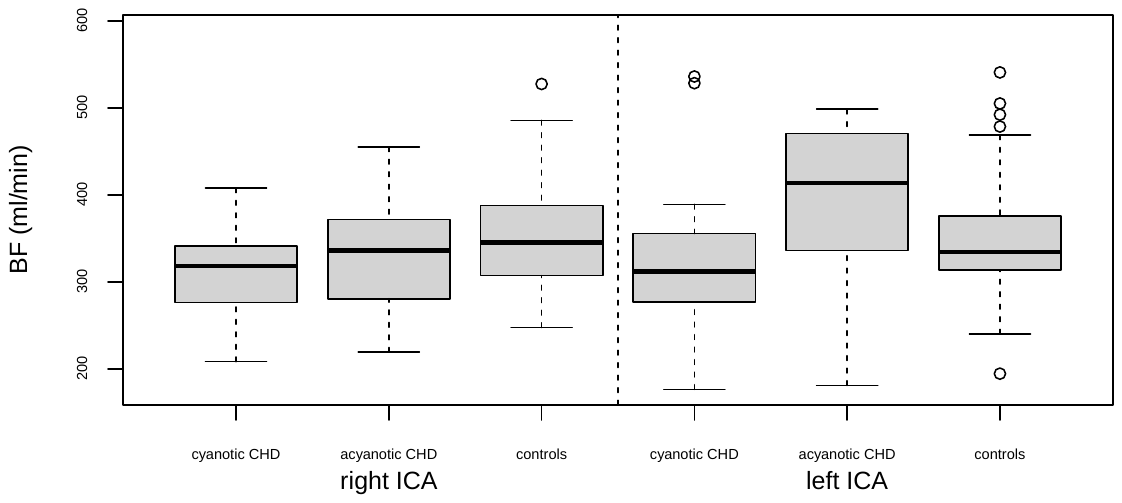**C** | 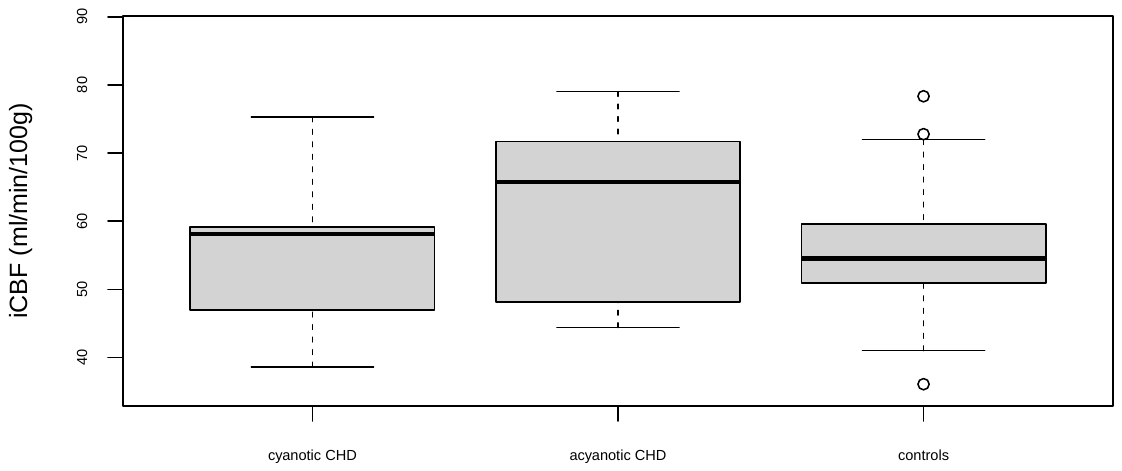**D** |
| 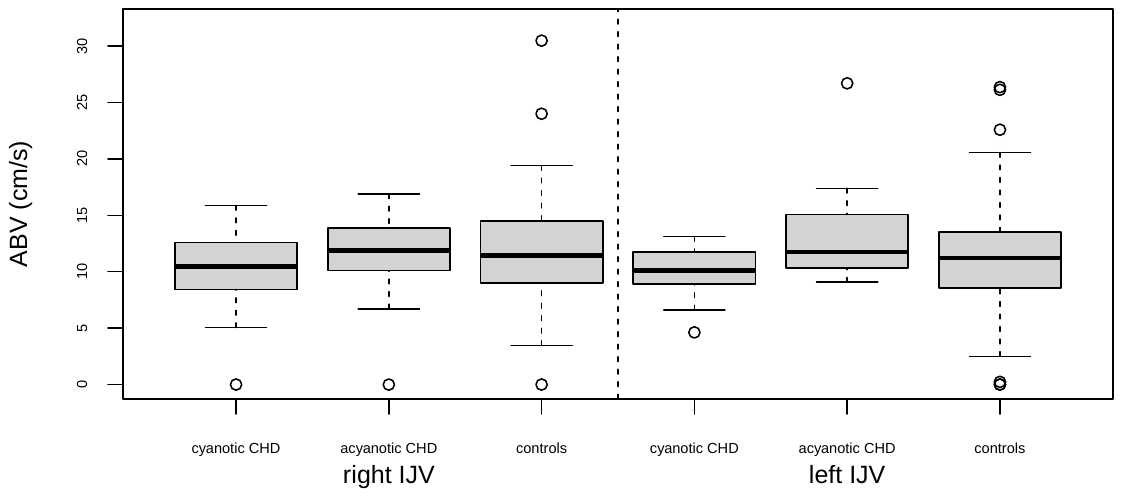**E** | 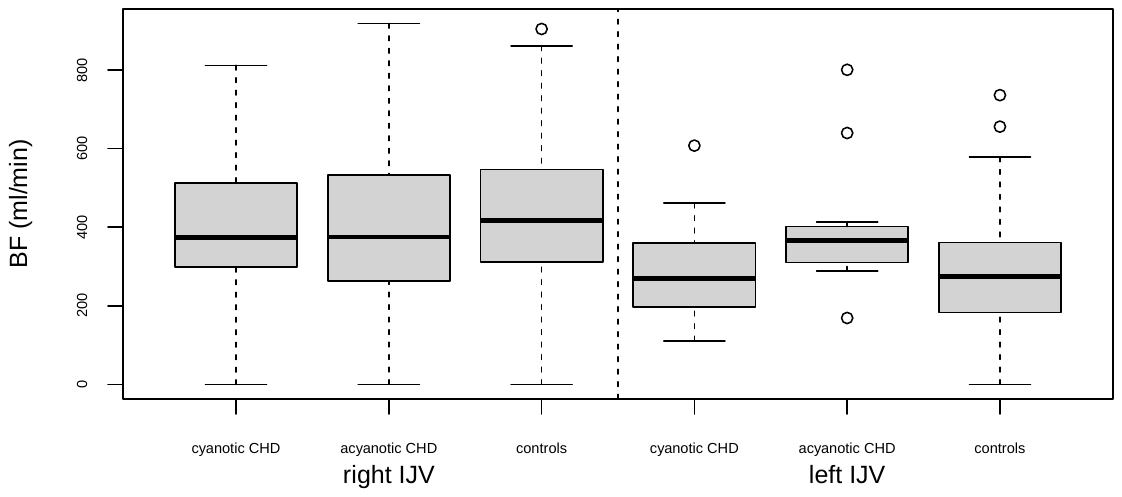**F** |

**Supplementary Figure 7. Boxplots of anterior cerebral hemodynamics among subgroups.**

Boxplots displaying all the cerebral hemodynamic parameters that were primary assessed in this study among patients with cCHD, aCHD and healthy controls.

cCHD=cyanotic congenital heart disease; aCHD=acyanotic congenital heart disease; ICA=internal carotid artery; ABV=average blood velocity; PBV=peak blood velocity; BF=blood flow; iCBF=indexed cerebral blood flow; IJV=internal jugular vein.
